# Epigenetic aging & embodying injustice: US My Body My Story and Multi-Ethnic Atherosclerosis Study

**DOI:** 10.1101/2023.12.13.23299930

**Authors:** Nancy Krieger, Christian Testa, Jarvis T. Chen, Nykesha Johnson, Sarah H. Watkins, Matthew Suderman, Andrew J. Simpkin, Kate Tilling, Pamela D. Waterman, Brent A. Coull, Immaculata De Vivo, George Davey Smith, Ana V. Diez Roux, Caroline Relton

## Abstract

**Importance:** Epigenetic accelerated aging is associated with exposure to social and economic adversity and may increase risk of premature morbidity and mortality. However, no studies have included measures of structural racism and few have compared estimates within or across the 1^st^ and 2^nd^ generation of epigenetic clocks (the latter additionally trained on phenotypic data).

**Objective:** To determine if accelerated epigenetic aging is associated with exposures to diverse measures of racialized, economic, and environmental injustice measured at different levels and time periods.

**Design:** Cross-sectional *My Body My Story Study* (MBMS; US, 2008-2010) and Exam 5 *Multi-Ethnic Atherosclerosis Study* (MESA; US, 2010-2012). MBMS DNA extraction: 2021; linkage of structural measures to MBMS and MESA: 2022.

**Setting:** MBMS recruited a random sample of US-born Black non-Hispanic (BNH) and white non-Hispanic (WNH) participants from 4 community health centers in Boston, MA. The MESA Exam 5 epigenetic component included 975 randomly selected US-born BNH, WNH, and Hispanic participants from four field sites: Baltimore, MD; Forsyth County, NC; New York City, NY; St. Paul, MN.

**Participants:** US-born persons (MBMS: 224 BNH, 69 WNH; MESA: 229 BNH, 555 WNH, 191 Hispanic).

**Main outcome and measures:** 10 epigenetic clocks (six 1^st^ generation; four 2^nd^ generation), computed using DNA methylation data (DNAm) from blood spots (MBMS; N = 293) and purified monocytes (MESA; N = 975).

**Results:** Among Black non-Hispanic MBMS participants, epigenetic age acceleration was associated with being born in a Jim Crow state by 0.14 standard deviations (95% confidence interval [CI] 0.00, 0.27) and with birth state conservatism (0.06, 95% CI 0.00, 0.05), pooling across all clocks, as was low parental education for both Black non-Hispanic and white non-Hispanic MBMS participants (respectively: 0.24, 95% CI 0.08, 0.39, and 0.27, 95% CI 0.03, 0.51. Adult impoverishment was positively associated with the pooled 2^nd^ generation clocks among the MESA participants (Black non-Hispanic: 0.06, 95% CI 0.01, 0.12; white non-Hispanic: 0.05, 95% CI 0.01, 0.08; Hispanic: 0.07, 95% CI 0.01, 0.14).

**Conclusions and Relevance:** Epigenetic accelerated aging may be one of the biological mechanisms linking exposure to racialized and economic injustice to well-documented inequities in premature morbidity and mortality.

**KEY POINTS:** **Question:** Is accelerated epigenetic aging associated with exposure to racialized, economic, and environmental injustice?

**Findings:** In the US cross-sectional *My Body My Story (MBMS; n = 263)* and *Multi-Ethnic Atherosclerosis Study (MESA, Exam 5; n = 1264)*), epigenetic accelerated aging was associated with Jim Crow birth state for MBMS Black non-Hispanic participants (by 0.14 standard deviations, 95% confidence interval 0.00, 0.27) and similarly with low parental education (MBMS: Black and white non-Hispanic participants) and adult impoverishment (MESA: Black and white non-Hispanic and Hispanic participants).

**Meaning:** Epigenetic accelerated aging may be a biological pathway for embodying racialized and economic injustice.

---

Identifying biological pathways by which societal injustice becomes embodied and expressed as population health inequities – that is, unjust, avoidable, and in principle preventable differences in health status across societal groups^1^ – is important, given implications for exposing accountability and preventing future harm.^1,2^ One set of potential pathways recently amenable to investigation in epidemiologic studies involves epigenetics, referring to diverse biological mechanisms that regulate gene expression and are heritable across cell generations.^3,4^ In 1940, the biologist Conrad Waddington (1906-1975) coined the term “epigenetics,” as part of his novel work investigating phenotypic development as a temporal path dependent process shaped by exposures affecting gene regulation and expression, canalization, and chance.^5^ Since then, epigenetic research has expanded to investigate, for example, such outcomes as morbidity, responsiveness to treatment, aging, and mortality.^3,4,6^

Of particular interest is the epigenetic mechanism of DNA methylation (DNAm), which entails the addition of a methyl group to cytosine, which chiefly occurs at cytosine-guanine dinucleotide (CpG) sites.^3,4,6^ Methylation at CpG-rich promoters tends to repress transcription, thereby reducing expression of the associated genes; methylation at other sites may have differential effects.^3,4,6^ DNAm is both dynamic and can be transmitted mitotically across cell generations.^6,7^ The discovery of age-related temporal patterning of DNAm led to creation, in 2013, of the first epigenetic “clocks” to estimate the biological age of an individual or a biological specimen,^7–10^ Comprising specified sets of DNAm sites, these epigenetic clocks “tick” in all cells and their descendants and yield estimates of “DNAm age” (or “epigenetic age”).^7,8^ The 1^st^ generation clocks used algorithms trained solely on chronological age; 2^nd^ generation clocks have additionally been trained using phenotypic data on health outcomes and exposures (e.g., cigarette smoking) and have also examined the “pace of aging” within individuals (**eTable 1**).^7,8,11,12^ Beyond estimating biological age, epigenetic clocks also enable the quantification of “epigenetic accelerated aging,” in essence DNAm age exceeding chronological age.^7–12^

Recent epidemiologic studies, summarized in several reviews, have reported that epigenetic accelerated aging is associated with not only increased risk of cardiometabolic disease, cancer, and earlier age at death but also well-known racialized and economic health inequities in these outcomes.^11–14^ Most of these epigenetic clock studies have focused on socioeconomic position, measured at the individual-level, with recent evidence (especially for 2^nd^ generation clocks^11,15,16^) showing stronger associations with early life as compared to adult deprivation.^11–16^ Several studies have documented associations with neighborhood poverty,^11–14,17^ air pollution,^11–14,18^ and adult self-reported experiences of racial discrimination^19,20^; none have included measures of structural racism or residential segregation.^21^

Guided by the ecosocial theory of disease distribution and its conceptualization of pathways of embodiment of societal injustice in relation to levels, lifecourse, and historical generation,^2^ we offer a novel analysis of associations between epigenetic accelerated aging among US Black non-Hispanic, white non-Hispanic, and Hispanic adults and exposure to Jim Crow birthplace (i.e., born in a US state in which white supremacy, before the passage of the 1964 and 1965 US Civil Rights Acts, was upheld by both state law and terror^2,22–25^) and current racialized economic segregation,^25–27^ along with exposure to self-reported experiences of racial discrimination, lifecourse socioeconomic position, and concurrent air pollution.

## METHODS

### Study Populations

We used data from two pre-existing US studies to conduct our analyses, and added new exposure data to both datasets. As pre-specified in our study protocol, the *My Body My Story* (MBMS) study^28,29^ comprised our “discovery” data (Boston, MA, 2008-2010), and a sub-set of the *Multi-Ethnic Study of Atherosclerosis* (MESA)^30,31^ served as a secondary data set (4 US cities, 2010-2012), one likely with different biases given differences in study design, older age range, and geographic heterogeneity.^32^

MBMS was designed by several current team members to investigate how racial discrimination and other social exposures affect risk of cardiovascular disease.^28,29^ As previously reported, the study recruited 1005 participants randomly selected from the patient rosters of four Boston community health centers between 2008 and 2010 who met study eligibility criteria: working age adults between age 35 and 64 years, born in the US, and self-identified as being either Black non-Hispanic or white non-Hispanic.^28,29^ Among the 1005 MBMS participants, 85% provided a finger prick blood sample on to filter paper (409 Black non-Hispanic; 466 white non-Hispanic), and within each racialized group, we determined that those who did vs. did not provide a blood spot did not differ by income level or self-reported experiences of racial discrimination. Participants’ blood spots were stored at −20°C and not analyzed until the present investigation.

MESA, a longitudinal cohort, obtained DNAm from a randomly selected subset of 1264 Exam 5 participants (2010-2012) drawn from the Baltimore, MD, Forsyth County, NC, New York City, NY and St. Paul, MN recruitment sites; they were age 55 to 94, and 582 were non-Hispanic white, 270 were non-Hispanic Black, 404 were Hispanic, and 10 were non-Hispanic of other race/ethnicities.^30,31^ At baseline (2000-2002), MESA recruited 6184 participants free of cardiovascular disease (CVD) ages 45 to 84 who self-identified as being white, African American, Hispanic, and Chinese, and were recruited from university-affiliated field centers in 6 US communities (the 4 named above plus Chicago, IL and Los Angeles, CA). MESA data for Exams 1-5 included social metrics analogous to those in MBMS.^30,31^ Per all MBMS participants being US-born, we focus on the 975 US-born MESA participants (229 Black non-Hispanic, 555 white non-Hispanic, 191 Hispanic).

### Epigenetic data

We extracted DNA from the MBMS blood spots in 2021, using the QIAamp DNA Investigator Kit for FTA and Guthrie cards, with samples randomized across 96 well plates, and measured DNAm using Illumina Infinium MethylationEPIC Beadchip; protocol details are provided in our preliminary study using the MBMS blood spots to demonstrate the feasibility and validity of analyzing specimens with as little as 40 ng DNA.^33^ Among the 875 MBMS blood spot specimens, 472 were judged suitable for DNA extraction; exclusions were due to an initial blood spot protocol problem affecting only the first community health center, whose members were predominantly white non-Hispanic. We then removed specimens from: (a) 50 participants with less than 40 ng DNA;^33^ (b) 96 participants with poor quality DNA extraction (as determined by high numbers of undetected probes on the EPIC BeadChip); and (c) 35 participants whose self-reported cis-gender identity did not match their chromosomal sex as predicted by *meffil* (an algorithm which relies on staining intensities for the X and Y chromosomes^34^); these latter specimens also displayed very little correlation between chronological age and epigenetic age. Thus, in total 293 MBMS participants (224 Black non-Hispanic and 69 white non-Hispanic) and 857,774 sites passed QC. Functional normalization included BeadArray as a fixed effect. Blood cell composition was estimated using a deconvolution algorithm,^35^ implemented in *meffil*, based on the “blood gse35069 complete” cell type reference.

DNAm for the MESA participants was measured in purified primary monocytes obtained from blood drawn in the morning after a 12-hour fast, using rigorous quality-controlled protocols.^36^ As described in detail in prior publications, epigenome-wide methylation was quantified using the llumina HumanMethylation450 BeadChip.^36^

We constructed the epigenetic clocks for all participants using code provided by the developers if available, or else using the formula in the original paper; values for GrimAge were calculated by uploading data to the GrimAge website (**eTable 1**). Among the 10 epigenetic clocks we computed, six were 1^st^ generation (Horvath; Hannum; Zhang Age; epiTOC; MiAge; DNAmTL) and four were 2^nd^ generation (Zhang Mortality; PhenoAge; Dunedin; Grim Age), with shared CpG sites more evident among the 1^st^ generation clocks (**eTables 1-3**). To address underlying genetic variability, we computed both surrogate variables and genetic principal components (PCs) (**eTextbox 1**) and included only the former in our models, as genetic PCs were available only for MESA and their inclusion did not alter results (**eFigure 1**).

### Exposure data

We conceptualized and operationalized the MBMS and MESA social metric exposure data in relation to type and level of exposure and time period of exposure (at time of birth, childhood, and adult) and stratified analyses by racialized groups, given their likely differences in exposure to and impacts of racialized injustice. Measures of exposure to racial injustice *at time of birth* included (with the latter two newly added): (a) born in a Jim Crow state^22,25,37^; (b) city of birth Index of Concentration at the Extremes (ICE) for racialized segregation^25–27^ (available for MBMS only); and (c) state of birth US state policy liberalism index, drawing on 148 policies enacted over 8 decades, including civil rights legislation.^38^ Validated adult measures of self-reported exposure to racial discrimination included, for MBMS, the Experiences of Discrimination scale,^25,39^ and, for MESA, the Major Discrimination Scale, restricted to unfair treatment attributed to race/ethnicity.^25,39^ Metrics for socioeconomic position prior to adulthood comprised self-reported highest level of educational attainment for the participants and for their parents.^28,29,32^ Additional self-report adult socioeconomic data at the individual and household levels pertained to: household income; household income per capita; household income to poverty ratio; employment status (and, if employed, occupational class (available in MBMS only); and housing tenure.^28,29,32^ Based on participants’ residential address at the time of the survey, we newly generated census tract level data for both the MBMS and MESA participants, using the American Community Survey 5-year estimates for 2008-2012^40^; these metrics included: composition by racialized group and the ICE for income, racialized, and racialized economic segregation, and housing tenure.^25–27^ Air pollution data for both MBMS and MESA were based on participants’ residential address at time of survey (details provided in **Table 1**). Missingness of the exposure data was typically under 5% for most variables (**Table 1**) and correlations among continuous variables from MBMS and MESA are respectively provided in **eFigures 2-3**.

**Table 1.**
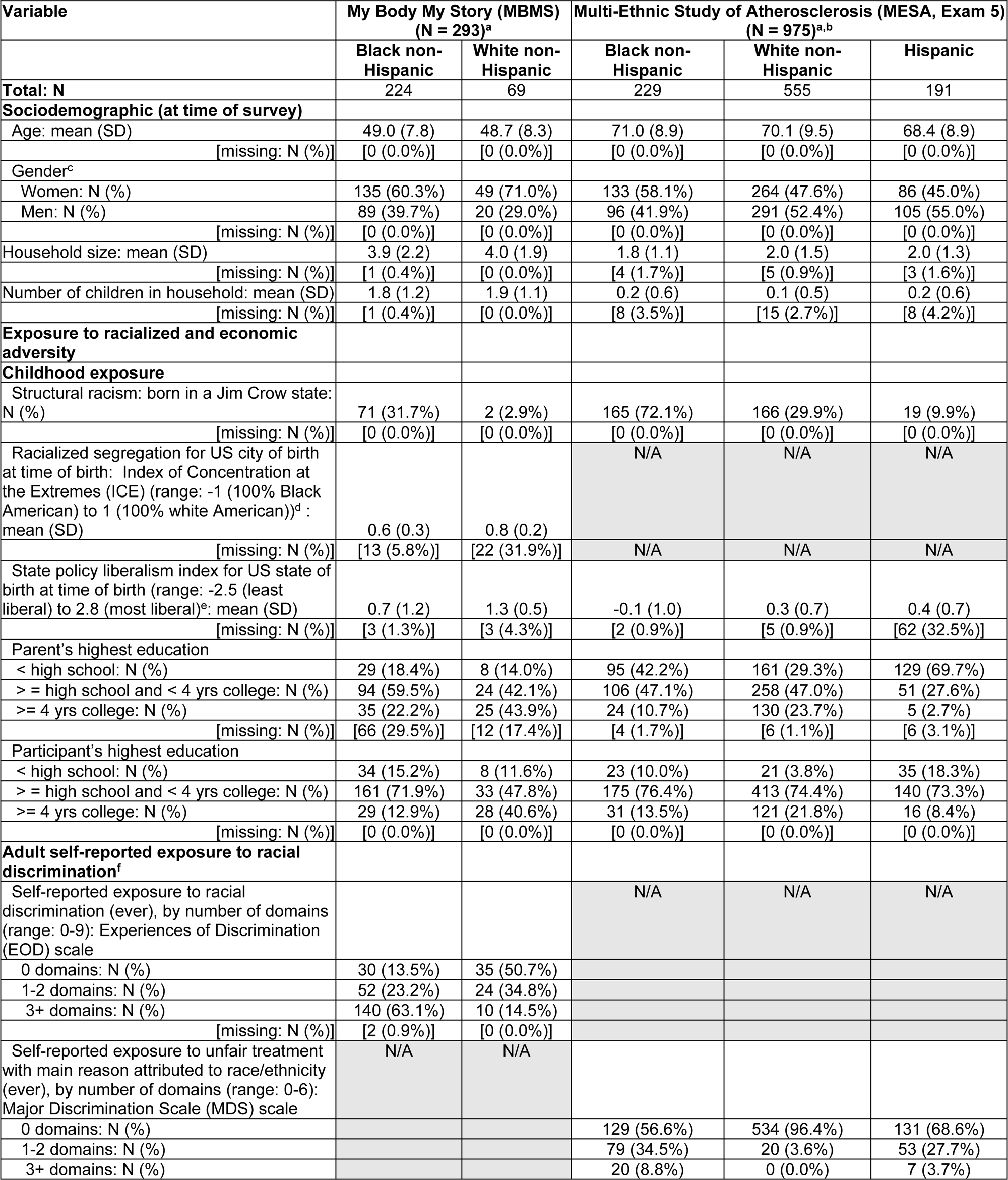
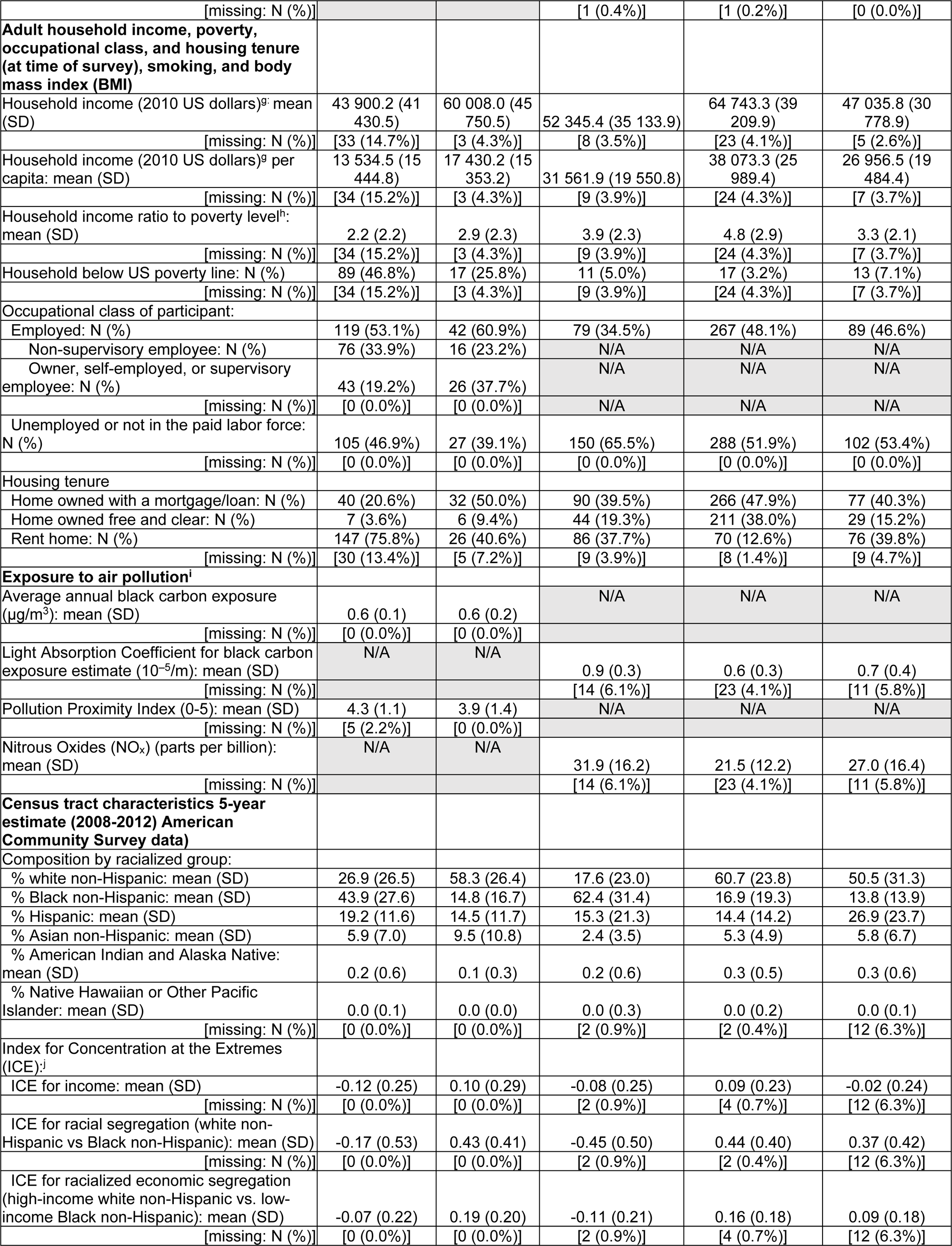
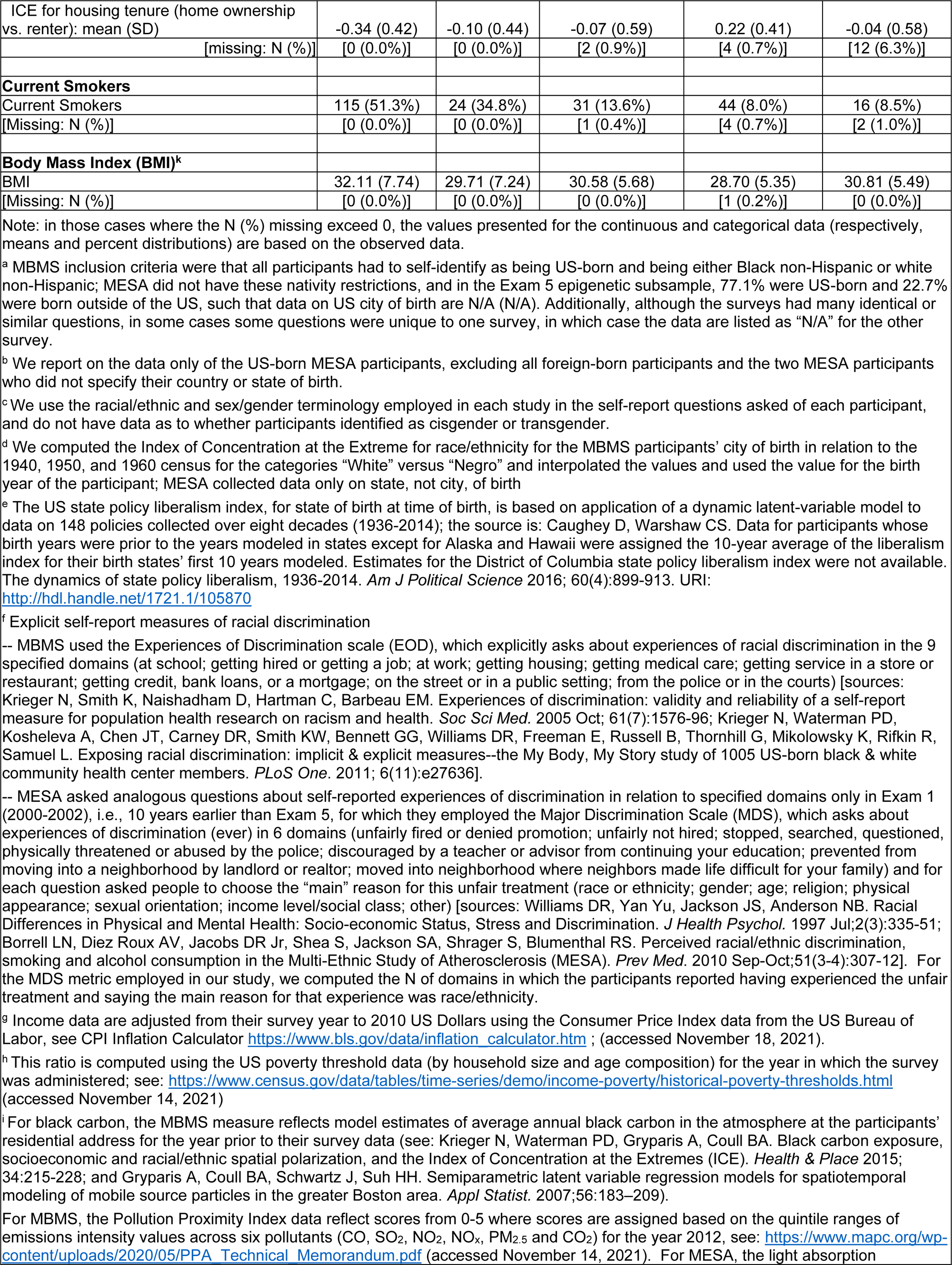
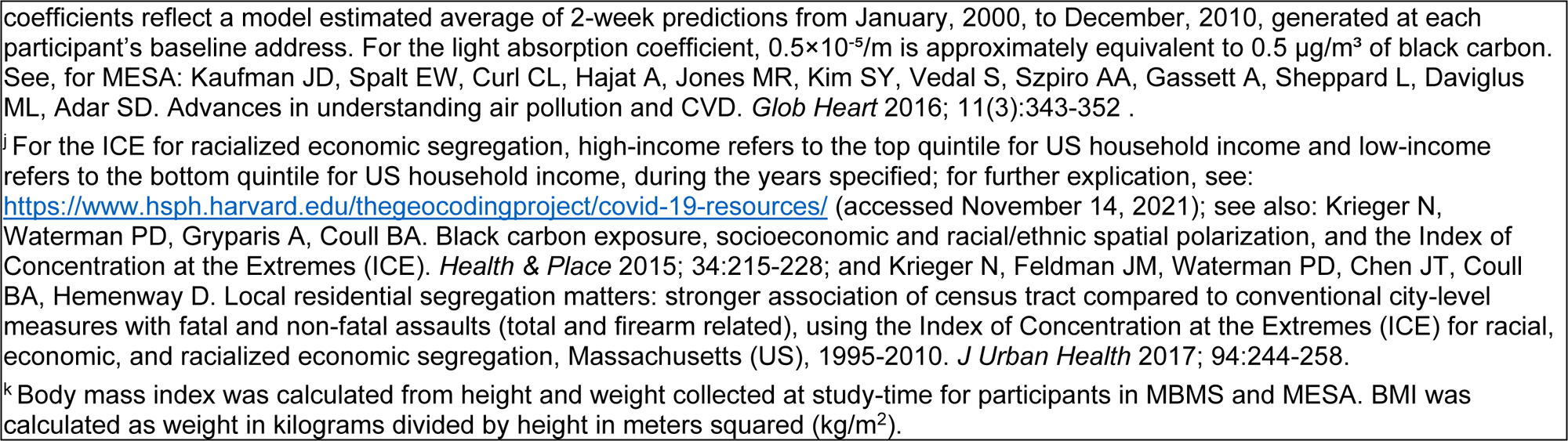
Exposure and covariate data for participants with epigenetic data: exposure to childhood and adult racialized and economic structural and household adversity, current sociodemographic characteristics, current adult exposure to air pollution, and current residential census tract social and economic context: *My Body My Story* study (MBMS; Boston, MA, 2008-2010; ages 35-64 years) and Multi-Ethnic Study of Atherosclerosis (MESA; 6 US sites, Exam 5 epigenetic subsample, 2010-2012; ages 55-94 years).

### Covariates

Additional covariates comprised self-reported data for: age; gender; and household size and number of children under age 18 (used to determine the household income to poverty ratio^41^).

### Statistical methods

Epigenetic clocks were regressed jointly on participants’ age, exposures to adversity (see Table 1), sex/gender, BMI, smoking, cell type proportions in their blood sample, and surrogate variables (see **eTextbox 1**). We fit separate models for each exposure-clock combination. Models were standardized to account for clocks’ scale differences. We employed crossed random-effects models^42^ with random intercepts by participant and random age slopes by clock to estimate average exposure effects assuming a common latent “epigenetic aging” construct observable across clocks. We used multiple imputation to account for missing data (**eTextbox 2**). We employed Bonferroni and False Discovery Rate correction for multiple comparisons testing (**eTextbox 2**).

## RESULTS

**Table 1** summarizes the distribution of the exposure data and covariates for the US-born MBMS study population (N = 293) and the US-born MESA study population (N = 975) with DNAm data, overall and stratified by racialized group (MBMS and MESA: Black non-Hispanic and white non-Hispanic; MESA only: Hispanic). MBMS participants on average were younger than MESA participants (means of 49.0 vs. 69.6 years) and more likely to self-identify as being women (62.8% vs. 51.4%). As expected, the Black non-Hispanic participants in both MBMS and MESA had the highest exposure to racial injustice at different levels and different points over the lifecourse (**Table 1**). They were most likely to be born in a Jim Crow state and/or a state with a low (least liberal) state policy liberalism index. As adults, they were also most likely to self-report exposure to racial discrimination in three or more domains and reside in segregated census tracts with high extreme concentrations of, respectively, Black, and low-income Black, non-Hispanic households. By contrast, the US-born white non-Hispanic MBMS and MESA participants were most likely to live in census tracts with high extreme concentrations of, respectively, white, and high-income white, non-Hispanic households (**Table 1**). Educational attainment was lower for participants’ parents compared to the participants, and in both groups was lowest for the Black non-Hispanic and Hispanic participants (**Table 1**). Similar patterns occurred for self-reported adult household income, poverty level, occupational class (if employed), and housing tenure, and also for the ICE metrics for income segregation and housing tenure. The MBMS and MESA DNAm raw age estimate and epigenetic accelerated aging estimates (detrended for chronological age) in relation to each specified exposure, stratified by racialized group (and, for MESA, by also stratified by nativity), and the cell type proportions, are presented for all 10 clocks in **eTable 4**.

In **Figures 1 through 4** we present standardized effect estimates for each clock, and also the pooled clock data, controlling for age, sex/gender, cell-type proportions, surrogate variables, smoking and BMI; in **eFigures 4-9**, we remove smoking and BMI.

**Figure 1.**
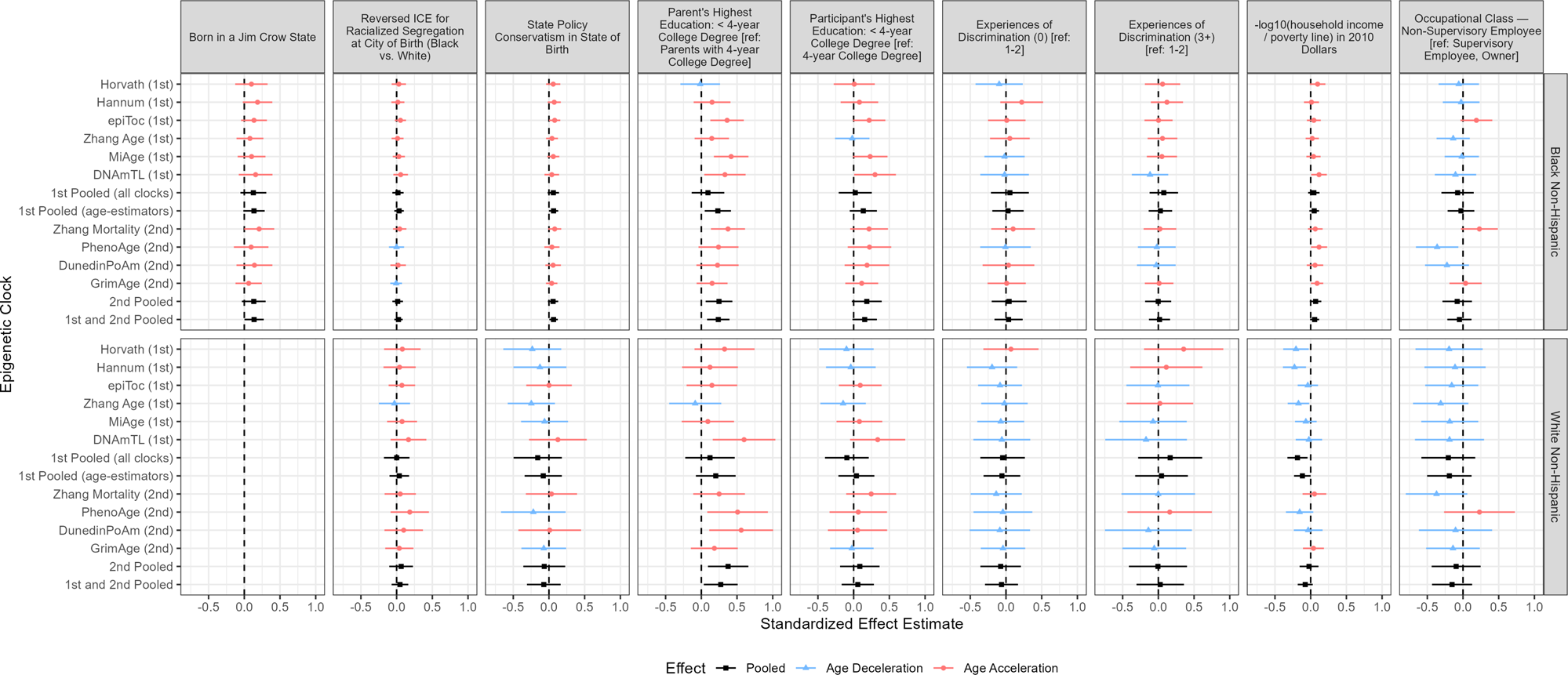
Standardized effect estimates and 95% confidence intervals by racialized group for early life adverse exposures (born in a Jim Crow state; reversed Index of Concentration at the Extremes for racialized segregation at city of birth; state policy conservatism in state of birth; parents’ education; participants’ education) and adult adverse exposures (Experiences of Discrimination; negative log household income ratio to poverty line; occupational class): MBMS participants. For interpretation, the effects of the 1^st^ generation age-estimator clocks can be viewed as how many additional years of aging (in units of standard deviations) are associated with the exposures – either in comparison to the reference category for categorical variables, or according to a 1-unit standard deviation increase in the exposure for the continuous measures. For reference, in the MBMS population, age had a standard deviation of 7.92, so a pooled effect estimate of 0.10 on the age-estimator clocks is comparable to an increase in aging of 0.79 years. Other clocks (as described in Supplemental Table 1) predict age-related measures such as mortality risk, mitotic divisions of stem cells, phenotypic outcomes, and telomere length, and therefore cannot be interpreted in units of additional years of aging. To ensure that the common effects of adverse exposures across clocks would be in the same direction, we: (a) reversed the direction of the DNAmTL clock (multiplied by negative one) because telomere length decreases with age, and (b) reversed the ICE measures compared to their conventional definitions, so that higher values represented more adverse exposures. All models controlled for the specified covariates (age, sex/gender, cell-type proportions, smoking, BMI, and surrogate variables). Because only 2 of the MBMS white non-Hispanic participants were born in a Jim Crow state (see Table 1), we do not report results for this group, given small numbers.

**Figure 2.**
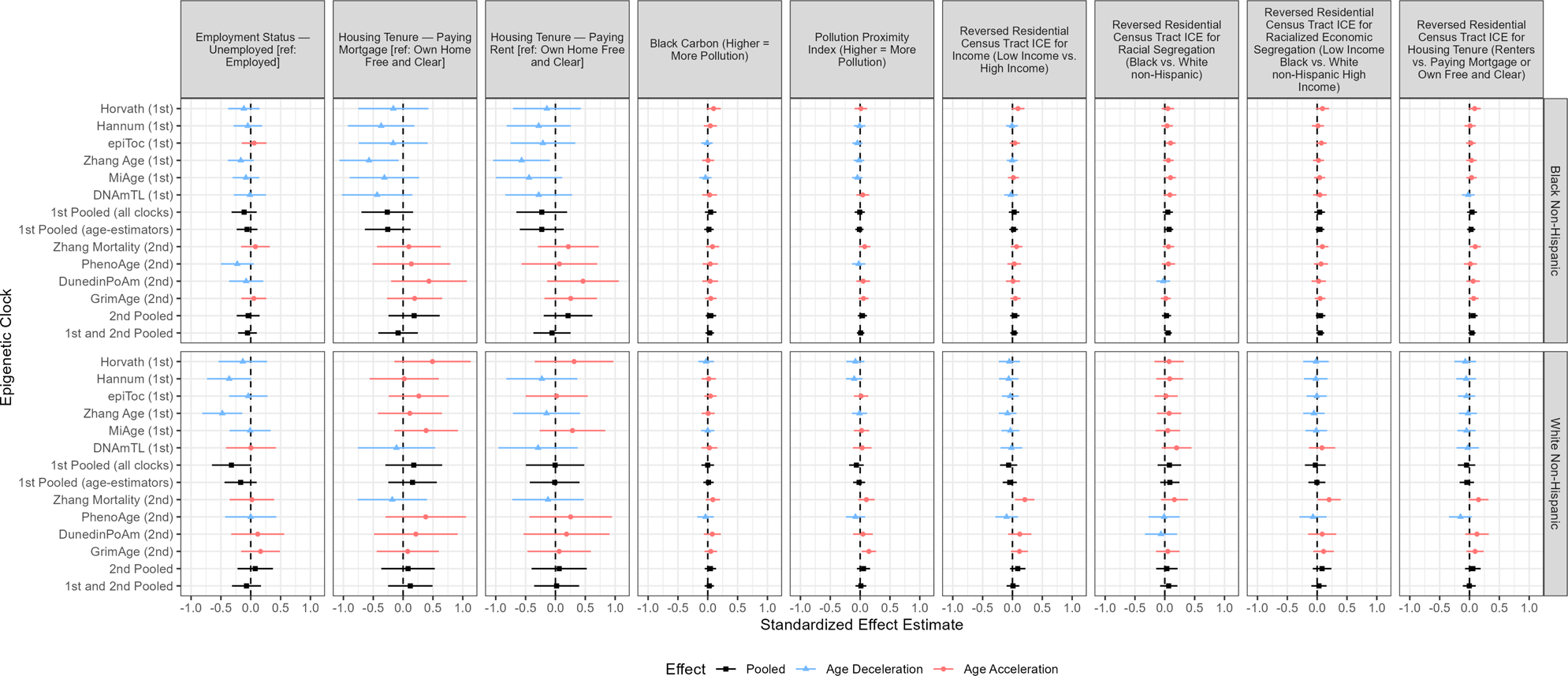
Standardized effect estimates and 95% confidence intervals by racialized group for adult individual-, household-, and area-based adverse exposures (employment status; housing tenure; black carbon; pollution proximity; reversed Index of Concentration at the Extremes for income, racial segregation, racialized economic segregation, and housing tenure): MBMS participants. Note that the Experiences of Discrimination measure refers specifically to those reported by participants as being due to their race, ethnicity, or color.

**Figure 3.**
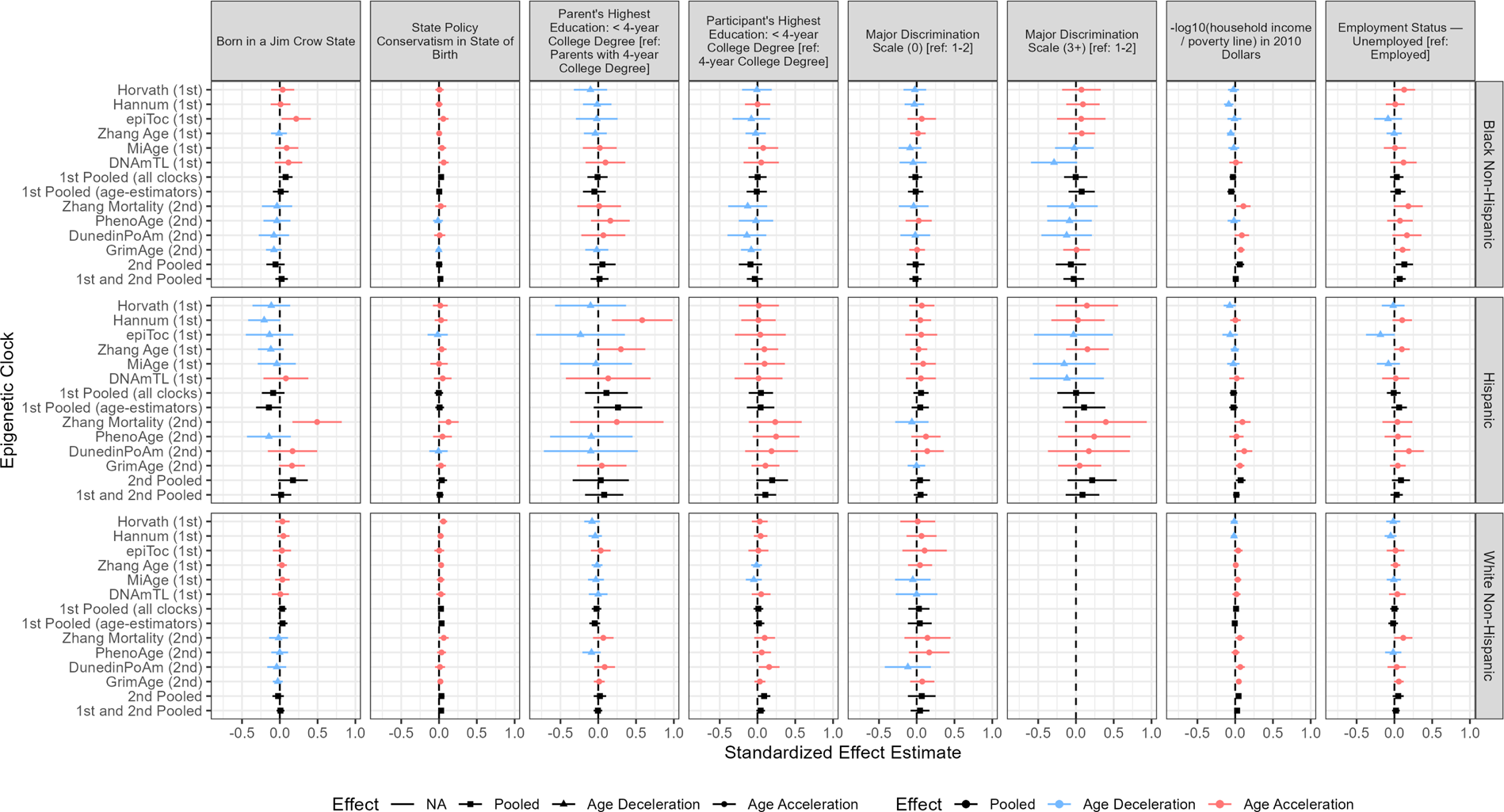
Standardized effect estimates and 95% confidence intervals by racialized group for early life adverse exposures (born in a Jim Crow state; state policy conservatism in state of birth; parents’ education; participants’ education) and adult adverse exposures (Major Discrimination scale (racialized); negative log household income ratio to poverty line; employment status): MESA participants Note: in the MESA population, age had a standard deviation of 9.36, so a pooled effect estimate of 0.10 on the age-estimator clocks is comparable to an increase in aging of 0.94 years. For the Major Discrimination Score (referring to discrimination reported by participants who attributed their reported unfair experiences as due to their race/ethnicity), no MESA white non-Hispanic participants had score >=3 (see Table 1), so there are no data to report for this category.

**Figure 4.**
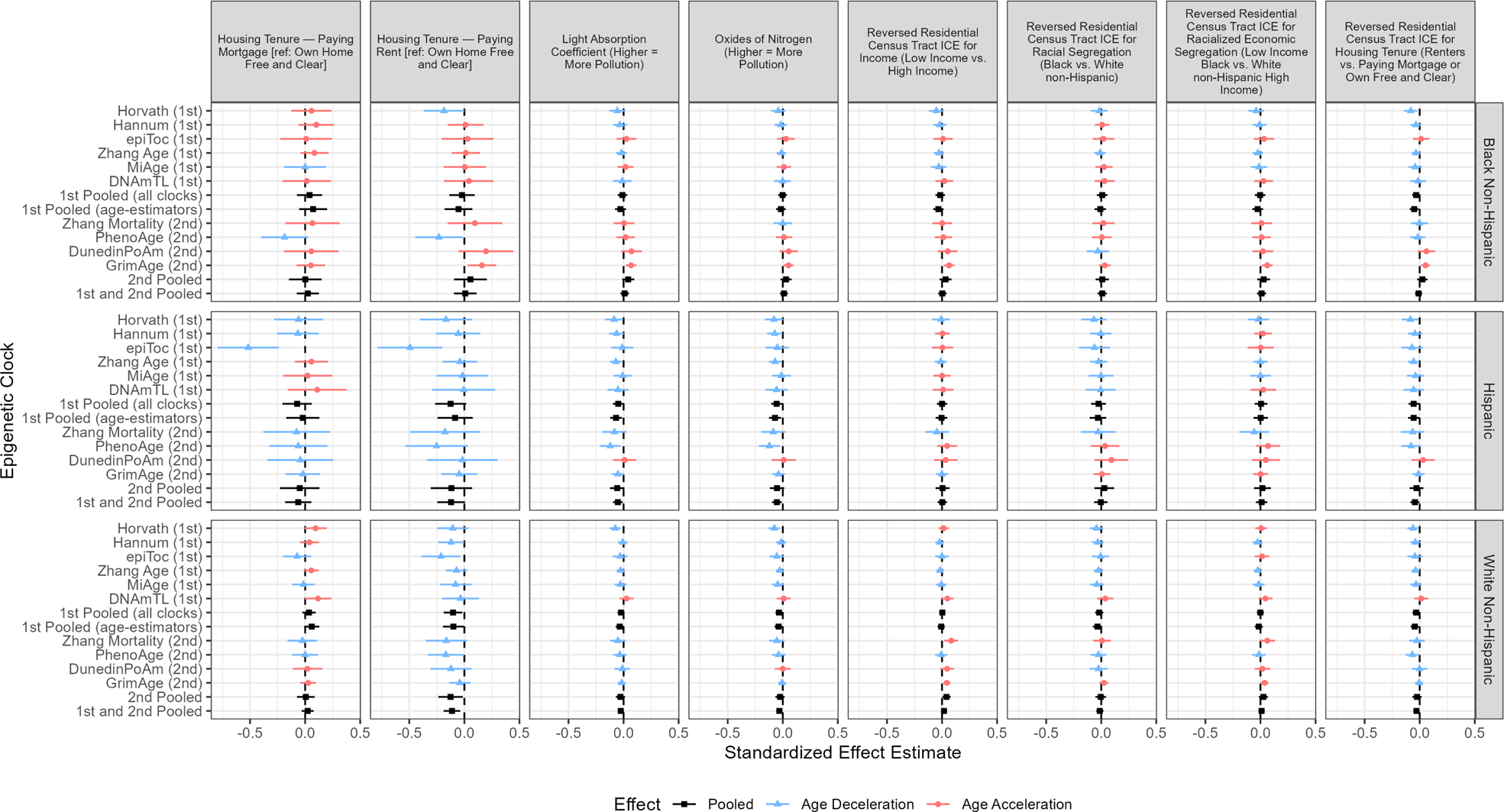
Standardized effect estimates and 95% confidence intervals by racialized group for adult household- and area-based adverse exposures (housing tenure; light absorption coefficient [air pollution]; oxides of nitrogen; reversed Index of Concentration at the Extremes for income, racial segregation, racialized economic segregation, and housing tenure): MESA participants

Our primary findings for MBMS were that we detected consistent patterns of epigenetic accelerated aging (with estimates whose 95% confidence intervals (CI) excluded 0), pooling across all clocks, for three social exposures: (a) Jim Crow birthplace and birth state conservatism for Black non-Hispanic participants only (0.14, 95% CI 0.00, 0.27 and 0.06, 95% CI 0.00, 0.12, respectively), and (b) low parental education, for both Black non-Hispanic and white non-Hispanic participants (respectively: 0.24, 95% CI 0.08, 0.39, and 0.27, 95% CI 0.03, 0.51); these latter estimates were similar or higher for the pooled 2^nd^ generation clocks for the Black non-Hispanic participants (0.25, 95% CI 0.06, 0.43), and the white non-Hispanic participants (0.38, 95% CI 0.09, 0.66).

Second, among the US-born MESA participants (most analogous to, albeit on average older than, the solely US-born MBMS participants), pooling across all clocks, we observed consistent patterns of accelerated epigenetic aging for the social exposures only for the white non-Hispanic participants for higher adult impoverishment (as measured by the negative log of adult household income ratio to the poverty line): 0.03, 95% CI 0.00, 0.05, and also birth state conservatism (0.03, 95% CI 0.00, 0.06). Adult impoverishment was also associated with epigenetic accelerated aging for the pooled 2^nd^ generation clocks for all three racialized groups (Black non-Hispanic: 0.06, 95% CI 0.01, 0.12; white non-Hispanic: 0.05, 95% CI 0.01, 0.08; Hispanic: 0.07, 95% CI 0.01, 0.14).

Correction for multiple comparisons testing (both Bonferroni and the less conservative False Discovery Rate) increased the p-values. Other social and environmental exposures did not exhibit consistent associations with epigenetic accelerated aging across clocks, among either MBMS or MESA participants. Additional analyses showed the expected associations of epigenetic accelerated aging with chronological age, BMI, and smoking (**eTable 5**) and also little difference in associations for exposures if analyses excluded smoking and BMI (**eFigures 4-9**).

## DISCUSSION

Our study offers intriguing novel evidence that exposure to Jim Crow at time of birth may be associated with epigenetic accelerated aging (pooling across all clocks) among US-born Black non-Hispanic working age adults, for whom being born in a Jim Crow state translated to age acceleration by 0.14 standard deviations (95% CI 0.00, 0.27) in MBMS, with evidence also of associations with birth state conservatism. It also adds to extant evidence that epigenetic accelerated aging, among working age or older adults, is associated with low parental education and adult impoverishment.^11–16^

Several study limitations merit consideration. First, despite having sample sizes on par with or larger than the handful of other epigenetic clock studies with data on exposure to racial injustice,^21^ our analytic sample sizes were relatively small, especially compared to much larger studies with more statistical power (with primarily European or US white populations) that have analyzed epigenetic clocks in relation to economic and pollution exposures,^12–14,18,43^ hence our focus on consistency of point estimates rather than statistical significance. Second, MBMS utilized frozen blood spots, a source validated for DNAm analyses,^44^ but MESA used DNAm obtained from purified monocytes. Additionally, as reported in our prior study demonstrating we could obtain results meeting rigorous quality control protocols using as little as 40ng of extracted DNA, albeit with decreasing statistical power with decreasing total weight of DNA.^33^ Third, for both MBMS and MESA, we used only DNAm from blood; yet, while DNAm patterns can vary by tissue, studies using DNAm measured in blood, including longitudinal analyses, have found robust associations between epigenetic accelerated aging and diverse health outcomes.^11–13^ Fourth, our observational study was not designed to yield causal effect estimates; generating data on associations with novel exposures, however, is a necessary first step. Foci for our next studies include EWAS analyses and investigating whether specific DNAm sites or epigenetic clocks mediate associations between the study exposures and selected health outcomes.

Study strengths include, first, conducting parallel analyses across two similar but not identical study populations, enhancing robustness of hypothesis testing, given likely different biases as described above.^32^ Under the assumption that the different clocks (variously trained on age, mortality, or phenotype data) reflect a latent construct of epigenetic aging, we computed standardized effect estimates to aid comparison of associations, singly and pooled. We also modeled age in our analysis using methods consistent with our prior work on identifying the correct way to model age in analyses of accelerated epigenetic aging.^21^

Supporting our view that the Jim Crow birthplace and parental education exposures provide suggestive signals of association, new research, using 2^nd^ generation clocks, has reported epigenetic accelerated aging to be associated with the economic shock of the 1930s US Depression^45^ and parents’ educational attainment.^16,46,47^ Inconsistencies in the MBMS and MESA results for Jim Crow place of birth merit further investigation and plausibly may reflect geographic variability in Jim Crow exposures, Jim Crow migration patterns, and exposure to racism in non-Jim Crow states.^22–25,37,48^ Other inconsistencies in the MBMS and MESA results may reflect differences in age of participants, cell types used for DNA extraction, and measurement on different Illumina beadchips (450K vs. EPIC). Suggesting that adult exposures may also matter is evidence linking epigenetic accelerated aging to adult economic deprivation and exposure to air pollution,^11–16,18,46,47,49^ with adults’ methylome shown to change in response to acute short-term exposures to air pollutants.^50^

Together, our findings, in conjunction with extant literature, support strengthening research to test the hypothesis that epigenetic accelerated aging may be one of the biological mechanisms underlying the well-documented elevated risk of premature morbidity and mortality, including for noncommunicable diseases, among social groups subjected to, vs. protected from, racialized and economic injustice.^2,11–16^ Future studies may benefit from inclusion of historically-informed measures of structural racialized and economic injustice contingent on the time period and geographic location where people are born, live, work, and bear and birth their children.

## Data Availability

This study relied on three sources of data, each of which is subject to distinct data sharing stipulations: (1) the non-public data from the My Body, My Story (MBMS) study; (2) the non-public data from the Multi-Ethnic Study of Atherosclerosis (MESA; data use agreement G638); and (3) the public de-identified data from the US Census, the American Community Survey, and the State Policy Liberalism Index. Descriptions of these data sharing stipulations and how to access these data is provided at: https://www.hsph.harvard.edu/nancy-krieger/data-sharing-resources/

https://reporter.nih.gov/search/T1qnoiJCukWRK0Mcvk-CJA/project-details/10363700

https://www.hsph.harvard.edu/nancy-krieger/data-sharing-resources/

https://github.com/DNAmAndAdversity/CensusData

https://github.com/DNAmAndAdversity/DNAmAndAdversity_public

https://zenodo.org/records/10256923

https://dataverse.harvard.edu/dataset.xhtml?persistentId=doi:10.7910/DVN/ZXZMJB

http://dx.doi.org/10.7910/DVN/ZXZMJB

https://internal.mesa-nhlbi.org/

https://github.com/shwatkins/epigenetic_clocks_mbms_mesa

## DECLARATION STATEMENTS

### 1) Ethics approval

The study protocol, involving use of both the MBMS and MESA data, was approved by the Harvard T.H. Chan School of Public Health Office of Human Research Administration (Protocol # IRB19-0524; June 10, 2019).

The original MBMS study protocol, implemented in accordance with the Helsinki Declaration of 1975, as revised in 2000, was approved by the Harvard School of Public Health Office of Human Research Administration (protocol #11950–127, which covered 3 of the 4 health centers through reciprocal IRB agreements), and was also separately approved by the fourth community health center’s Institutional Review Board. All participants provided written informed consent.

Information regarding the MESA protocols and their IRB approvals and other information, is available at: www.mesa-nhlbi.org.

### 2) Data sharing

This study (NIH Grant number R01MD014304) relied on three sources of data, each of which is subject to distinct data sharing stipulations: (1) the non-public data from the “My Body, My Story” (MBMS) study; (2) the non-public data from the Multi-Ethnic Study of Atherosclerosis (MESA; data use agreement G638); and (3) the public de-identified data from the US Census, the American Community Survey, and the State Policy Liberalism Index. We provide descriptions of these data sharing stipulations and access to these data below; this information is also available at: https://www.hsph.harvard.edu/nancy-krieger/data-sharing-resources/

- ICE metrics relating to racial composition, income distribution, and housing tenure that were derived from sources in the public domain i.e. the US Census and the American Community Survey are available at the census tract level now on GitHub.
- Code used to construct the variables as well as code used for our analyses are also available on GitHub and here.
- The State Policy Liberalism Index data used in our study is also publicly available and can be obtained from the Harvard Dataverse. Reference: Caughey, Devin; Warshaw, Christopher, 2014, “The Dynamics of State Policy Liberalism, 1936-2014”, http://dx.doi.org/10.7910/DVN/ZXZMJB Dataverse [Distributor] V1 [Version].
- De-identified data from the *My Body My Story* study used for this project will be made available only for purposes approved by the study PI, as stipulated by the study’s informed consent protocol. The application form to obtain these data will be made available via this website after completion of this project in late Fall 2024.
- Data from the Multi-Ethnic Study of Atherosclerosis (MESA) must be obtained directly from the MESA website via their application protocol.
- The scripts to create the epigenetic clocks constructed from raw methylation data are available on GitHub.

### 3) Author contributions

- NK led conceptualization of the study, co-designed the analyses, wrote the first draft, and co-led obtaining funds for the research project.
- CT accessed the electronic public use data and generated the study variables derived from these data, co-designed and conducted the analyses, produced the tables and figures, drafted text for the methods section, contributed to interpreting results, and prepared the study materials to be shared via the data repository (data dictionary; software code).
- JTC co-designed and supervised the analyses and contributed to interpreting results.
- NJ assisted with data analysis, producing the figures, and submitting the study materials to be shared via the data repository, and contributed to interpreting results.
- PDW facilitated finalizing all human subject approvals and data use agreements and also the data transfer of the MBMS epigenetic data from HSPH to Bristol, geocoded the place of birth data, extracted the historical census data from PDF files, and contributed to interpreting results.
- CR co-led obtaining funds for the research project and contributed to designing the analyses and interpreting results.
- SW and MS contributed to designing the analyses, prepared the analytic epigenetic clock data from the raw MBMS epigenetic data and the raw MESA epigenetic data, prepared the epigenetic Surrogate Variables and the genetic PC variables, and contributed to interpreting the results.
- AS, BC, KT, and GDS contributed to designing the analyses and interpreting the results.
- IDV led and supervised the assays to extract epigenetic data from the MBMS blood spots and contributed to designing the analyses and interpreting the results.
- ADR facilitated interpretation of the MESA data and contributed to designing the analyses and interpreting the study results.
- All co-authors provided critical intellectual content to and approved the submitted manuscript.

### 4) Funding

The work for this study was supported by the National Institutes of Health, National Institute of Minority and Health Disparities [grant number grant R01MD014304 to N.K. and C.R., as MPIs]. Additionally, GDS, CR, KT, MS, SHW work within the MRC Integrative Epidemiology Unit at the University of Bristol, which is supported by the Medical Research Council (MC_UU_00011/1,3, & 5 and MC_UU-00032/1). The funders had no role in study design, data collection and analysis, decision to publish, or preparation of the manuscript.

#### Prior funding supporting our work

a. The prior MBMS study was funded by the National Institutes of Health/National Institute on Aging (R01AG027122) and generation of the black carbon data for this study was supported by a pilot grant from the Harvard NIEHS Center (P30 ES000002).
b. No additional grant funding to MESA was obtained for the conduct of this study whose Data Use Agreement (G638) was approved by MESA (on 11/22/2019) to use pre-existing MESA data.

Support for generating these pre-existing MESA data is provided by contracts 75N92020D00001, HHSN268201500003I, N01-HC-95159, 75N92020D00005, N01-HC-95160, 75N92020D00002, N01-HC-95161, 75N92020D00003, N01-HC-95162, 75N92020D00006, N01-HC-95163, 75N92020D00004, N01-HC-95164, 75N92020D00007, N01-HC-95165, N01-HC-95166, N01-HC-95167, N01-HC-95168, N01-HC-95169, UL1-TR-000040, UL1-TR-001079, UL1-TR-001420, UL1-TR-001881, and DK063491. The MESA Epigenomics & Transcriptomics Studies were funded by NIH grants 1R01HL101250, 1RF1AG054474, R01HL126477, R01DK101921, and R01HL135009.

This publication was developed under the Science to Achieve Results (STAR) research assistance agreements, No. RD831697 (MESA Air) and RD-83830001 (MESA Air Next Stage), awarded by the U.S Environmental Protection Agency (EPA). It has not been formally reviewed by the EPA. The views expressed in this document are solely those of the authors and the EPA does not endorse any products or commercial services mentioned in this publication.

Funding for MESA SHARe genotyping was provided by NHLBI Contract N02-HL-6-4278. Genotyping was performed at the Broad Institute of Harvard and MIT (Boston, Massachusetts, USA) and at Affymetrix (Santa Clara, California, USA) using the Affymetrix Genome-Wide Human SNP Array 6.0

## 5) Acknowledgements

We thank, with written permission:

1. Linda V. Nguyen (Department of Medicine, Brigham and Women’s Hospital and Harvard Medical School, Boston, MA, USA; May 1, 2023) for her contributions to extracting the DNA from the MBMS blood spots and preparing it for shipment to the Bristol team members for analysis;
2. Emily Wright, PhD (Harvard TH Chan School of Public Health; May 1, 2023), for her assistance, when a doctoral student, with linking area-based data to the MBMS study participant records; and
3. Karen Ho (May 17, 2023), Louise Falk (May 18, 2023), Sophie Fitzgerald (May 24, 2023), and Catherine Slack (May 19, 2023) for their role in generating the MBMS Illumina data.

We are extremely grateful to all the MBMS participants who took part in the original MBMS study, the staff at the four participating community health centers, and the researchers who conducted participant recruitment and interviews. We are likewise grateful to all the MESA participants and investigators whose data and dedicated work enabled us to have incorporate MESA data into our research project, including Amanda Gassett, MS for providing us with the air pollution dataset used from MESA Air (written permission: July 20, 2023); a full list of participating MESA investigators and institutions can be found at: http://www.mesa-nhlbi.org. This paper has been reviewed and approved by the MESA Publications and Presentations Committee (Manuscript G-893, approved October 20, 2023).

## SUPPLEMENTARY MATERIALS

### 1) Supplementary eTables

**eTable 1.**
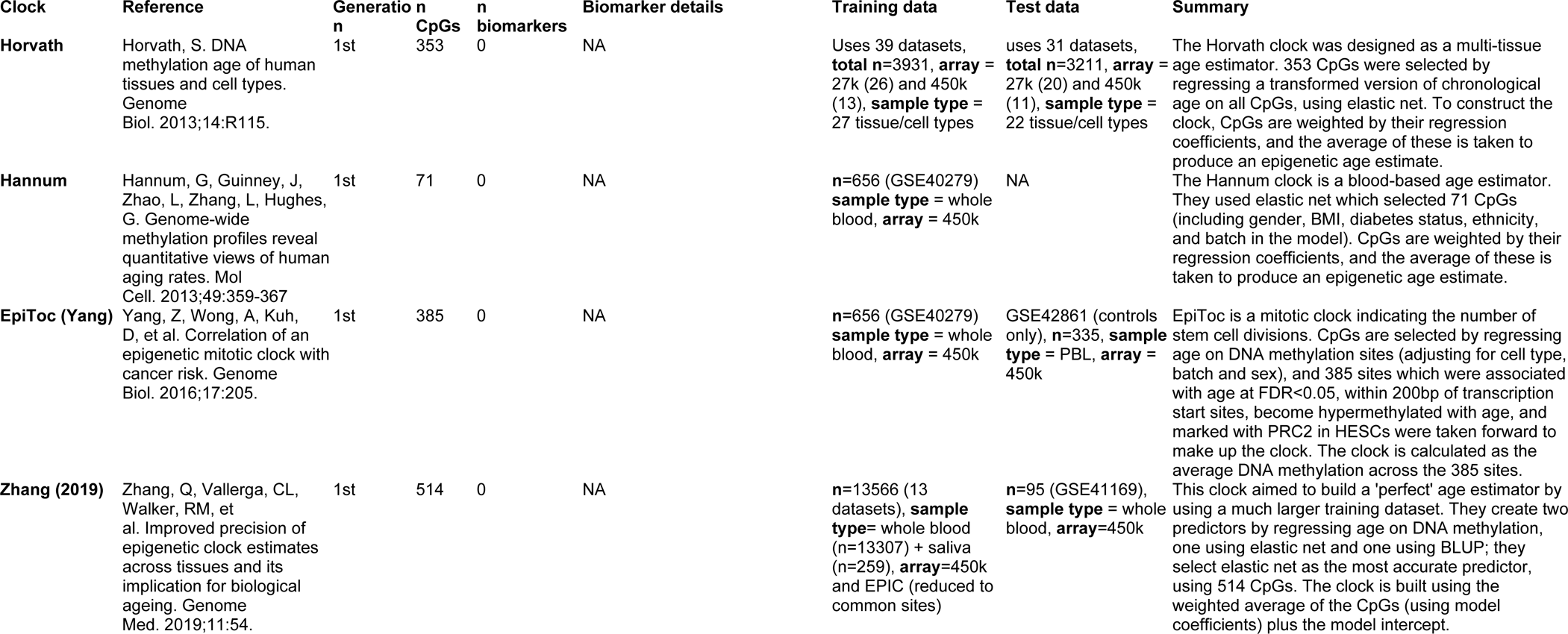
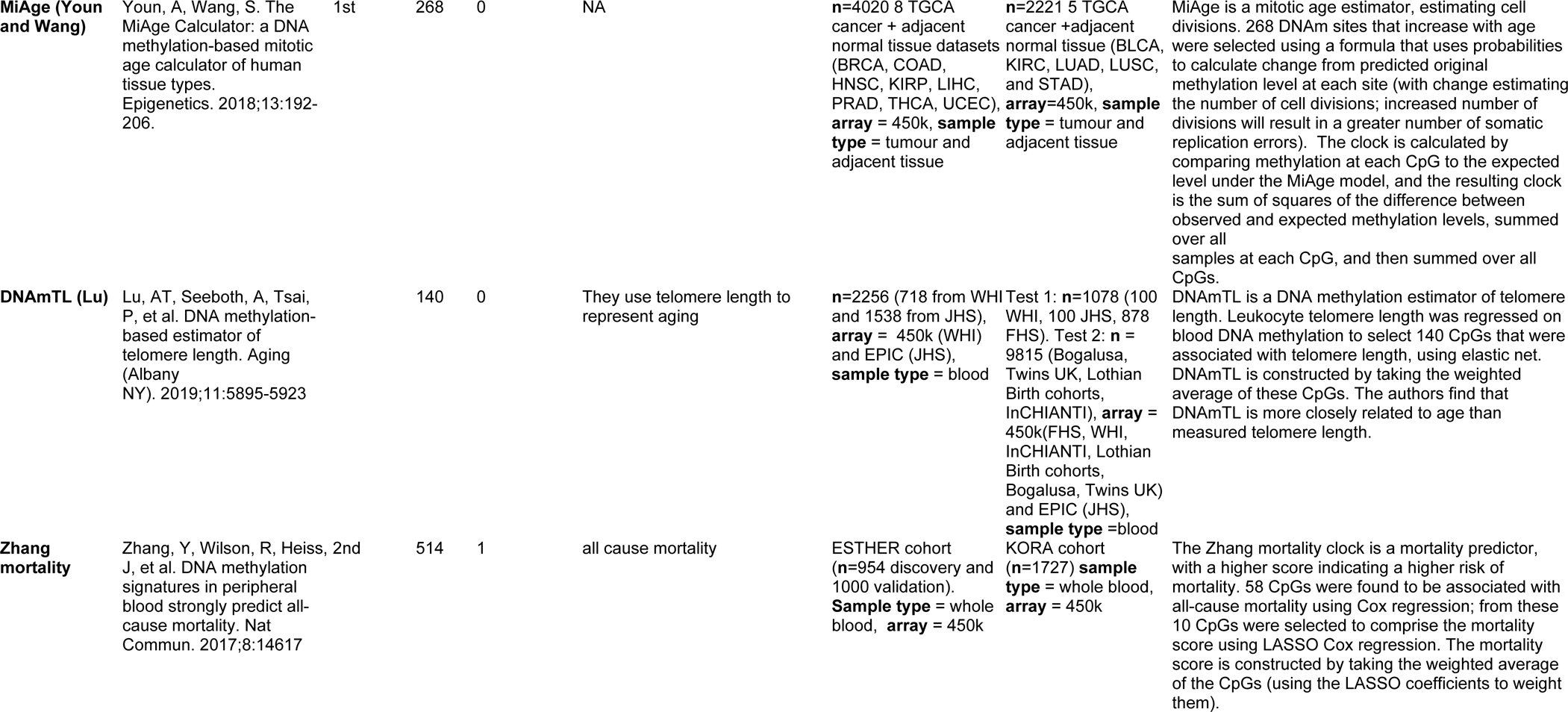
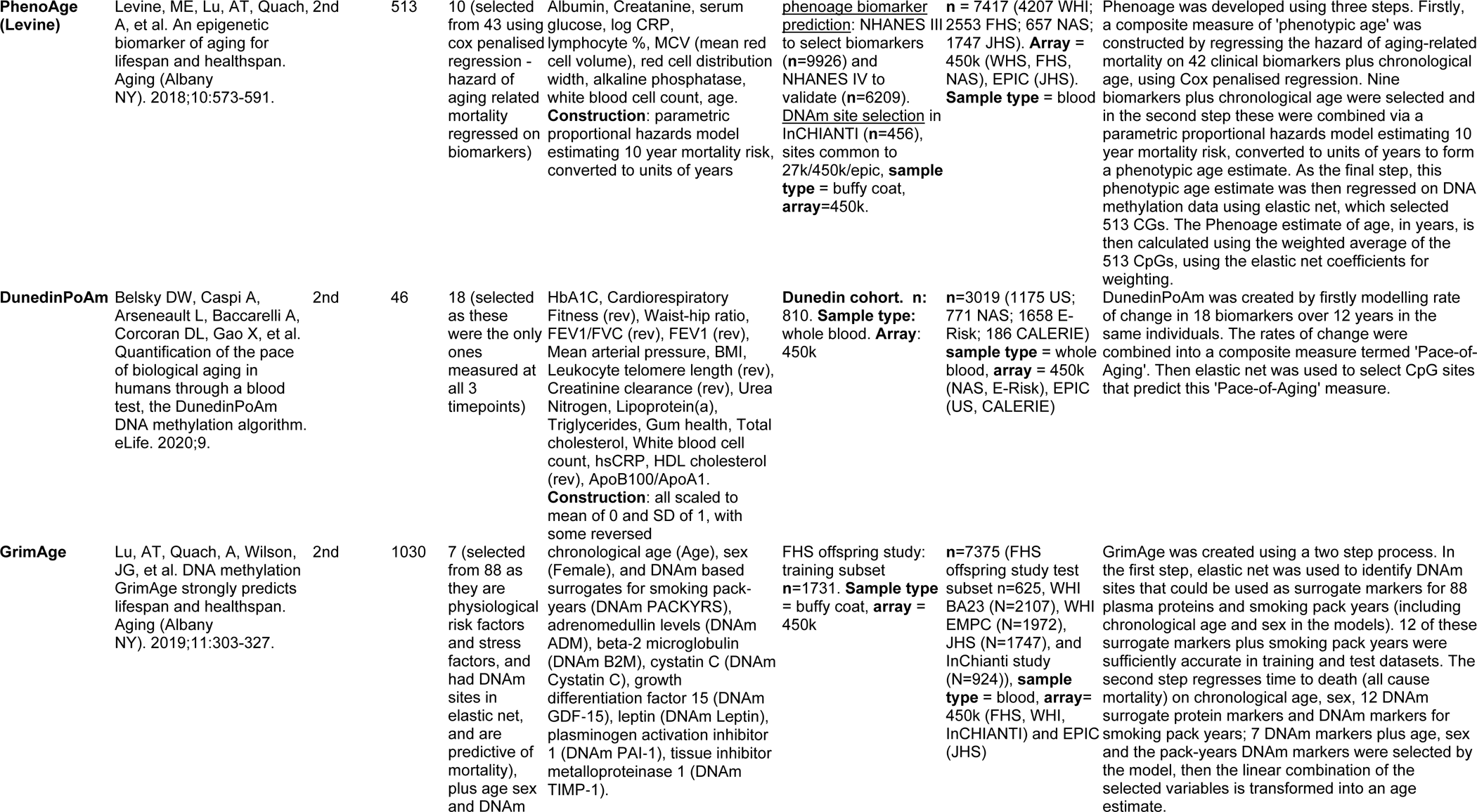
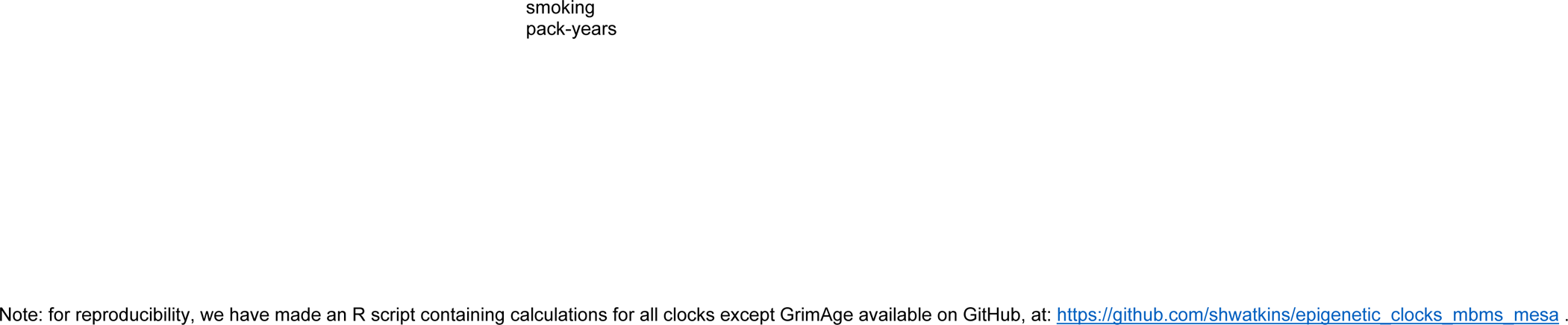
Primary citations for and descriptions of the 10 epigenetic clocks employed in this study.

**eTable 2.** Software dependency citations (using citations specified by each source).

|  |
| --- |
| 1. R Core Team (2022). R: A language and environment for statistical computing. R Foundation for Statistical Computing, Vienna, Austria. URL https://www.R-project.org/. |
| 2. Ou J (2021). `_colorBlindness`: Safe Color Set for Color Blindness. R package version 0.1.9, < https://CRAN.R-project.org/package=colorBlindness >. |
| 3. Firke S (2021). `_janitor`: Simple Tools for Examining and Cleaning Dirty Data . R package version 2.1.0, < https://CRAN.R-project.org/package=janitor >. |
| 4. Bache S, Wickham H (2022). `_magrittr`: A Forward-Pipe Operator for R . R package version 2.0.3, < https://CRAN.R-project.org/package=magrittr >. |
| 5. Hadley Wickham. `testthat`: Get Started with Testing. *The R Journal*, vol. 3, no. 1, pp. 5--10, 2011 |
| 6. Wickham H, Averick M, Bryan J, Chang W, McGowan LD, François R, Grolemond G, Hayes A, Henry L, Hester J, Kuhn M, Pedersen TL, Miller E, Bache SM, Müller K, Ooms J, Robinson D, Seidel DP, Spinu V, Takahashi K, Vaughan D, Wilke C, Woo K, Yutani H (2019). “Welcome to the tidyverse.” *Journal of Open Source Software*, *4*(43), 1686. doi:10.21105/joss.01686 < https://doi.org/10.21105/joss.01686 >. |
| 7. Robinson D, Hayes A, Couch S (2022). `_broom`: Convert Statistical Objects into Tidy Tibbles. R package version 1.0.2, < https://CRAN.R-project.org/package=broom >. |
| 8. Bolker B, Robinson D (2022). `_broom.mixed`: Tidying Methods for Mixed Models. R package version 0.2.9.4, < https://CRAN.R-project.org/package=broom.mixed >. |
| 9. Stef van Buuren, Karin Groothuis-Oudshoorn (2011). `mice`: Multivariate Imputation by Chained Equations in R. *Journal of Statistical Software*, 45(3), 1-67. DOI 10.18637/jss.v045.i03. |
| 10. Douglas Bates, Martin Maechler, Ben Bolker, Steve Walker (2015). Fitting Linear Mixed-Effects Models Using lme4. *Journal of Statistical Software*, 67(1), 1-48. doi:10.18637/jss.v067.i01. |
| 11. Müller K (2020). `_here`: A Simpler Way to Find Your Files. R package version 1.0.1, < https://CRAN.R-project.org/package=here >. |
| 12. Schauburger P, Walker A (2022). `_openxlsx`: Read, Write and Edit xlsx Files. R package version 4.2.5.1, < https://CRAN.R-project.org/package=openxlsx >. |
| 13. Hadley Wickham, Jennifer Bryan. R Packages (2e). O'Reilly; Available from: https://r-pkgs.org/ |
| 14. JJ Allaire and Yihui Xie and Jonathan McPherson and Javier Luraschi and Kevin Ushey and Aron Atkins and Hadley Wickham and Joe Cheng and Winston Chang and Richard Iannone (2022). `rmarkdown`: Dynamic Documents for R. R package version 2.19. URL https://rmarkdown.rstudio.com. |
| 15. Leek JT, Johnson WE, Parker HS, Fertig EJ, Jaffe AE, Zhang Y, Storey JD, Torres LC (2023). `sva`: *Surrogate Variable Analysis*. R package version 3.48.0. |

**eTable 3.** Number of overlapping CpG sites shared between clocks.

|  | Horvath<br>(1st) | Hannum<br>(1st) | epiToc<br>(1st) | Zhang Age<br>(1st) | MiAge<br>(1st) | DNAmTL<br>(2nd) | Zhang Mortality<br>(2nd) | PhenoAge<br>(2nd) | Dunedin<br>(2nd) |
| --- | --- | --- | --- | --- | --- | --- | --- | --- | --- |
| Horvath (1st) | 353 | 6 | 1 | 11 | 0 | 0 | 0 | 41 | 0 |
| Hannum (1st) | 6 | 71 | 0 | 30 | 0 | 5 | 0 | 6 | 0 |
| epiToc (1st) | 1 | 0 | 385 | 0 | 2 | 0 | 0 | 1 | 0 |
| Zhang Age (1st) | 11 | 30 | 0 | 515 | 1 | 6 | 0 | 16 | 0 |
| MiAge (1st) | 0 | 0 | 2 | 1 | 268 | 0 | 0 | 0 | 0 |
| DNAmTL (2nd) | 0 | 5 | 0 | 6 | 0 | 140 | 0 | 1 | 0 |
| Zhang Mortality (2nd) | 0 | 0 | 0 | 0 | 0 | 0 | 10 | 0 | 1 |
| PhenoAge (2nd) | 41 | 6 | 1 | 16 | 0 | 1 | 0 | 514 | 0 |
| Dunedin (2nd) | 0 | 0 | 0 | 0 | 0 | 0 | 1 | 0 | 46 |

**eTable 4.**
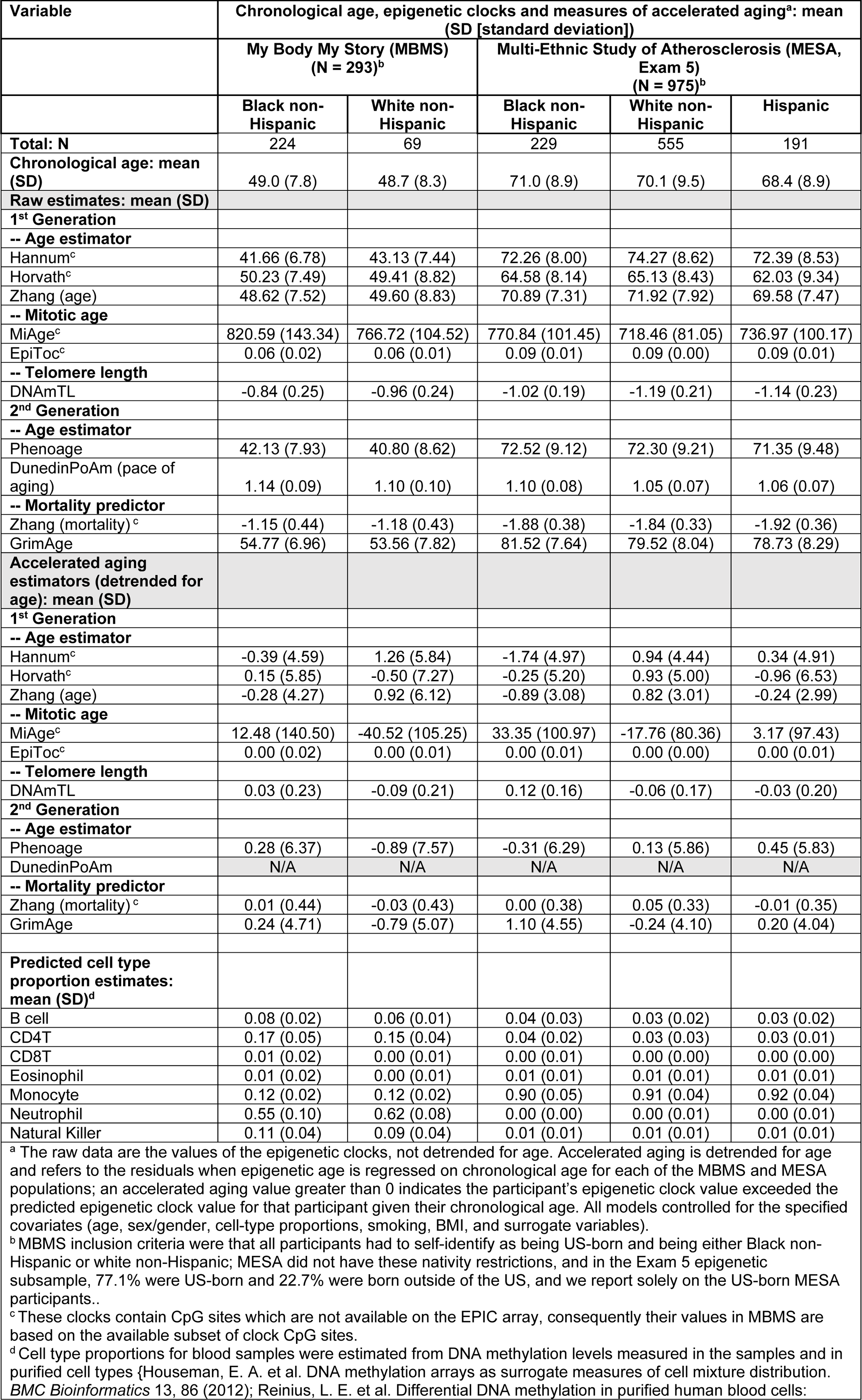
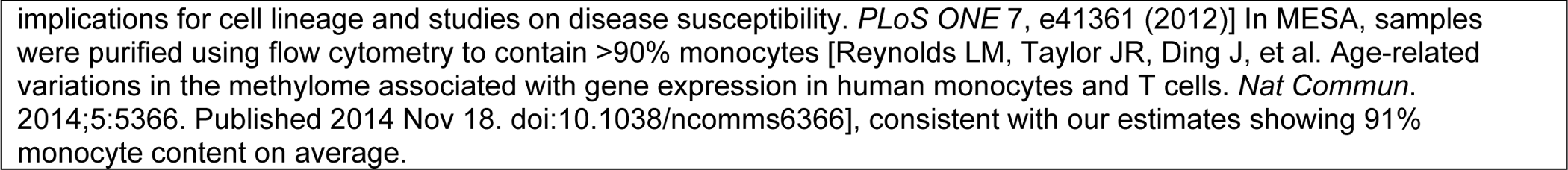
Chronological age, epigenetic clock and accelerated aging data, and cell type proportion covariates: *My Body My Story* study (MBMS; Boston, MA, 2008-2010; ages 35-64 years) and Multi-Ethnic Study of Atherosclerosis (MESA; 6 US sites, Exam 5 epigenetic subsample, 2010-2012; ages 55-94 years, US-born).

**eTable 5.**
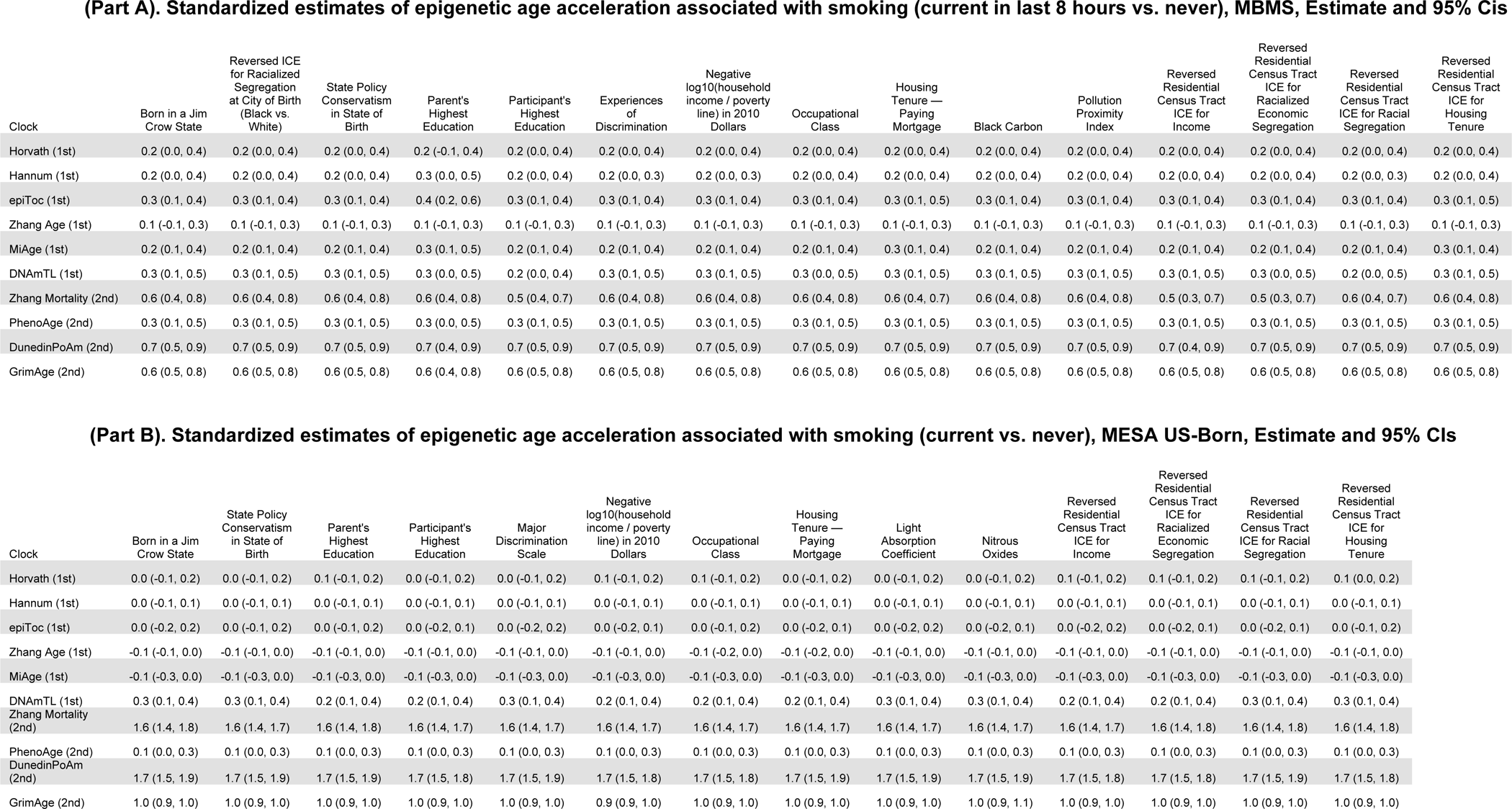
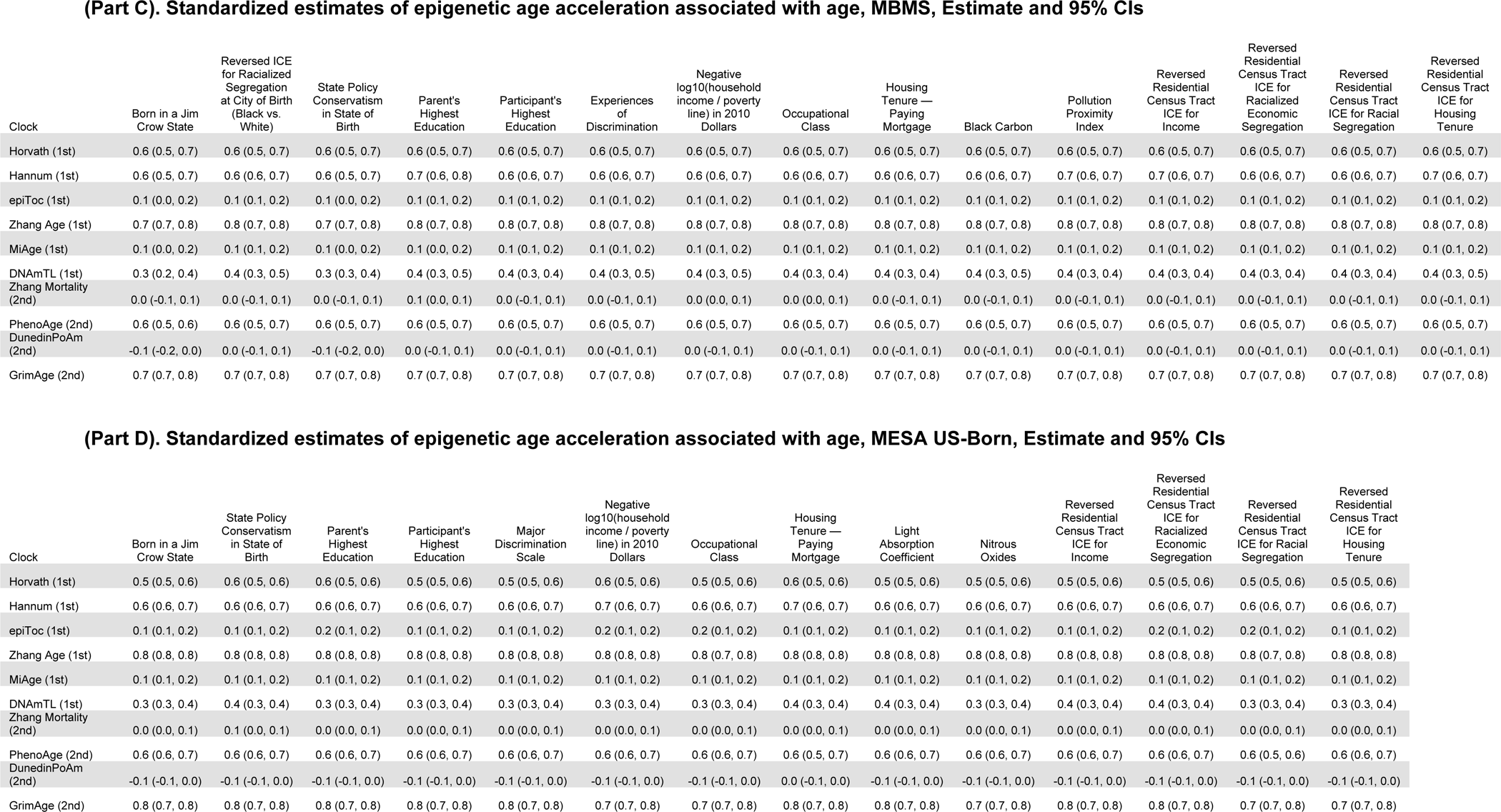
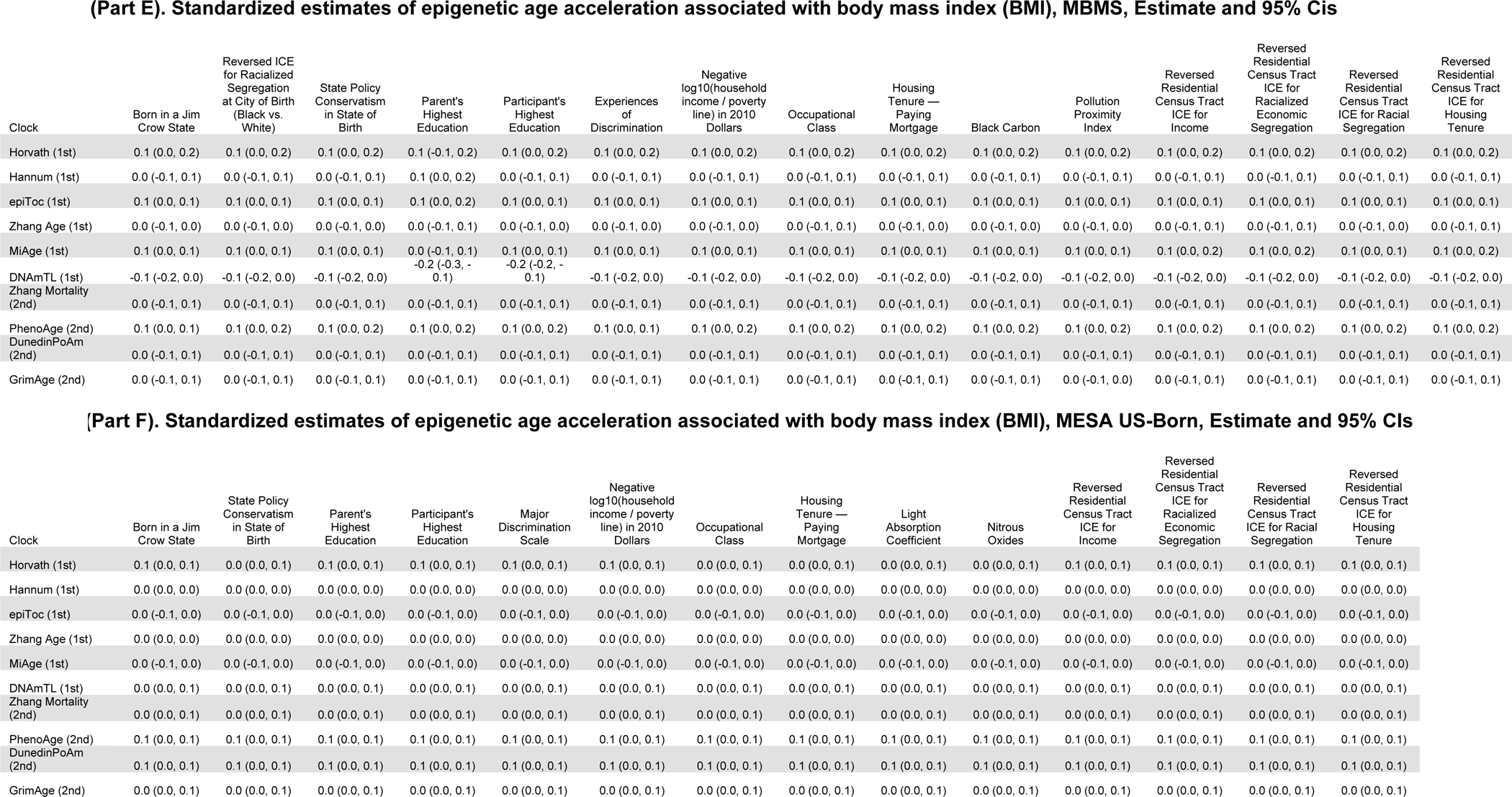
(Parts A-F). Standardized estimates of epigenetic age acceleration associated with smoking, age, and body mass index (BMI), for MBMS and MESA US-Born, Estimate and 95% CIs; each model includes the specified exposure, age, sex/gender, cell-type proportions, smoking, BMI, and surrogate variables.

| Clock | Born in a Jim Crow State | Reversed ICE for Racialized Segregation at City of Birth (Black vs. White) | State Policy Conservatism in State of Birth | Parent's Highest Education | Participant's Highest Education | Experiences of Discrimination | Negative log10(household income / poverty line) in 2010 Dollars | Occupational Class | Housing Tenure — Paying Mortgage | Black Carbon | Pollution Proximity Index | Reversed Residential Census Tract ICE for Income | Reversed Residential Census Tract ICE for Racialized Economic Segregation | Reversed Residential Census Tract ICE for Racial Segregation | Reversed Residential Census Tract ICE for Housing Tenure |
| --- | --- | --- | --- | --- | --- | --- | --- | --- | --- | --- | --- | --- | --- | --- | --- |
| Horvath (1st) | 0.2 (0.0, 0.4) | 0.2 (0.0, 0.4) | 0.2 (0.0, 0.4) | 0.2 (-0.1, 0.4) | 0.2 (0.0, 0.4) | 0.2 (0.0, 0.4) | 0.2 (0.0, 0.4) | 0.2 (0.0, 0.4) | 0.2 (0.0, 0.4) | 0.2 (0.0, 0.4) | 0.2 (0.0, 0.4) | 0.2 (0.0, 0.4) | 0.2 (0.0, 0.4) | 0.2 (0.0, 0.4) | 0.2 (0.0, 0.4) |
| Hannum (1st) | 0.2 (0.0, 0.4) | 0.2 (0.0, 0.4) | 0.2 (0.0, 0.4) | 0.3 (0.0, 0.5) | 0.2 (0.0, 0.4) | 0.2 (0.0, 0.3) | 0.2 (0.0, 0.3) | 0.2 (0.0, 0.4) | 0.2 (0.0, 0.4) | 0.2 (0.0, 0.4) | 0.2 (0.0, 0.4) | 0.2 (0.0, 0.4) | 0.2 (0.0, 0.4) | 0.2 (0.0, 0.3) | 0.2 (0.0, 0.4) |
| epiToc (1st) | 0.3 (0.1, 0.4) | 0.3 (0.1, 0.4) | 0.3 (0.1, 0.4) | 0.4 (0.2, 0.6) | 0.3 (0.1, 0.4) | 0.3 (0.1, 0.4) | 0.3 (0.1, 0.4) | 0.3 (0.1, 0.4) | 0.3 (0.1, 0.5) | 0.3 (0.1, 0.4) | 0.3 (0.1, 0.4) | 0.3 (0.1, 0.4) | 0.3 (0.1, 0.4) | 0.3 (0.1, 0.4) | 0.3 (0.1, 0.5) |
| Zhang Age (1st) | 0.1 (-0.1, 0.3) | 0.1 (-0.1, 0.3) | 0.1 (-0.1, 0.3) | 0.1 (-0.1, 0.3) | 0.1 (-0.1, 0.3) | 0.1 (-0.1, 0.3) | 0.1 (-0.1, 0.3) | 0.1 (-0.1, 0.3) | 0.1 (-0.1, 0.3) | 0.1 (-0.1, 0.3) | 0.1 (-0.1, 0.3) | 0.1 (-0.1, 0.3) | 0.1 (-0.1, 0.3) | 0.1 (-0.1, 0.3) | 0.1 (-0.1, 0.3) |
| MiAge (1st) | 0.2 (0.1, 0.4) | 0.2 (0.1, 0.4) | 0.2 (0.1, 0.4) | 0.3 (0.1, 0.5) | 0.2 (0.1, 0.4) | 0.2 (0.1, 0.4) | 0.2 (0.1, 0.4) | 0.2 (0.1, 0.4) | 0.3 (0.1, 0.4) | 0.2 (0.1, 0.4) | 0.2 (0.1, 0.4) | 0.2 (0.1, 0.4) | 0.2 (0.1, 0.4) | 0.2 (0.1, 0.4) | 0.3 (0.1, 0.4) |
| DNAmTL (1st) | 0.3 (0.1, 0.5) | 0.3 (0.1, 0.5) | 0.3 (0.1, 0.5) | 0.3 (0.0, 0.5) | 0.2 (0.0, 0.4) | 0.3 (0.1, 0.5) | 0.3 (0.1, 0.5) | 0.3 (0.0, 0.5) | 0.3 (0.1, 0.5) | 0.3 (0.1, 0.5) | 0.3 (0.1, 0.5) | 0.3 (0.1, 0.5) | 0.3 (0.0, 0.5) | 0.2 (0.0, 0.5) | 0.3 (0.1, 0.5) |
| Zhang Mortality (2nd) | 0.6 (0.4, 0.8) | 0.6 (0.4, 0.8) | 0.6 (0.4, 0.8) | 0.6 (0.4, 0.8) | 0.5 (0.4, 0.7) | 0.6 (0.4, 0.8) | 0.6 (0.4, 0.8) | 0.6 (0.4, 0.8) | 0.6 (0.4, 0.7) | 0.6 (0.4, 0.8) | 0.6 (0.4, 0.8) | 0.5 (0.3, 0.7) | 0.5 (0.3, 0.7) | 0.6 (0.4, 0.7) | 0.6 (0.4, 0.8) |
| PhenoAge (2nd) | 0.3 (0.1, 0.5) | 0.3 (0.1, 0.5) | 0.3 (0.1, 0.5) | 0.3 (0.0, 0.5) | 0.3 (0.1, 0.5) | 0.3 (0.1, 0.5) | 0.3 (0.1, 0.5) | 0.3 (0.1, 0.5) | 0.3 (0.1, 0.5) | 0.3 (0.1, 0.5) | 0.3 (0.1, 0.5) | 0.3 (0.1, 0.5) | 0.3 (0.1, 0.5) | 0.3 (0.1, 0.5) | 0.3 (0.1, 0.5) |
| DunedinPoAm (2nd) | 0.7 (0.5, 0.9) | 0.7 (0.5, 0.9) | 0.7 (0.5, 0.9) | 0.7 (0.4, 0.9) | 0.7 (0.5, 0.9) | 0.7 (0.5, 0.9) | 0.7 (0.5, 0.9) | 0.7 (0.5, 0.9) | 0.7 (0.5, 0.9) | 0.7 (0.5, 0.9) | 0.7 (0.5, 0.9) | 0.7 (0.4, 0.9) | 0.7 (0.5, 0.9) | 0.7 (0.5, 0.9) | 0.7 (0.5, 0.9) |
| GrimAge (2nd) | 0.6 (0.5, 0.8) | 0.6 (0.5, 0.8) | 0.6 (0.5, 0.8) | 0.6 (0.4, 0.8) | 0.6 (0.5, 0.8) | 0.6 (0.5, 0.8) | 0.6 (0.5, 0.8) | 0.6 (0.5, 0.8) | 0.6 (0.5, 0.8) | 0.6 (0.5, 0.8) | 0.6 (0.5, 0.8) | 0.6 (0.5, 0.8) | 0.6 (0.5, 0.8) | 0.6 (0.5, 0.8) | 0.6 (0.5, 0.8) |

| Clock | Born in a Jim Crow State | State Policy Conservatism in State of Birth | Parent's Highest Education | Participant's Highest Education | Major Discrimination Scale | Negative log10(household income / poverty line) in 2010 Dollars | Occupational Class | Housing Tenure — Paying Mortgage | Light Absorption Coefficient | Nitrous Oxides | Reversed Residential Census Tract ICE for Income | Reversed Residential Census Tract ICE for Racialized Economic Segregation | Reversed Residential Census Tract ICE for Racial Segregation | Reversed Residential Census Tract ICE for Housing Tenure |
| --- | --- | --- | --- | --- | --- | --- | --- | --- | --- | --- | --- | --- | --- | --- |
| Horvath (1st) | 0.0 (-0.1, 0.2) | 0.0 (-0.1, 0.2) | 0.1 (-0.1, 0.2) | 0.0 (-0.1, 0.2) | 0.0 (-0.1, 0.2) | 0.1 (-0.1, 0.2) | 0.1 (-0.1, 0.2) | 0.0 (-0.1, 0.2) | 0.0 (-0.1, 0.2) | 0.0 (-0.1, 0.2) | 0.1 (-0.1, 0.2) | 0.1 (-0.1, 0.2) | 0.1 (-0.1, 0.2) | 0.1 (0.0, 0.2) |
| Hannum (1st) | 0.0 (-0.1, 0.1) | 0.0 (-0.1, 0.1) | 0.0 (-0.1, 0.1) | 0.0 (-0.1, 0.1) | 0.0 (-0.1, 0.1) | 0.0 (-0.1, 0.1) | 0.0 (-0.1, 0.1) | 0.0 (-0.1, 0.1) | 0.0 (-0.1, 0.1) | 0.0 (-0.1, 0.1) | 0.0 (-0.1, 0.1) | 0.0 (-0.1, 0.1) | 0.0 (-0.1, 0.1) | 0.0 (-0.1, 0.1) |
| epiToc (1st) | 0.0 (-0.2, 0.2) | 0.0 (-0.1, 0.2) | 0.0 (-0.1, 0.2) | 0.0 (-0.2, 0.1) | 0.0 (-0.2, 0.2) | 0.0 (-0.2, 0.1) | 0.0 (-0.1, 0.2) | 0.0 (-0.2, 0.1) | 0.0 (-0.2, 0.2) | 0.0 (-0.2, 0.1) | 0.0 (-0.2, 0.2) | 0.0 (-0.2, 0.1) | 0.0 (-0.2, 0.1) | 0.0 (-0.1, 0.2) |
| Zhang Age (1st) | -0.1 (-0.1, 0.0) | -0.1 (-0.1, 0.0) | -0.1 (-0.1, 0.0) | -0.1 (-0.1, 0.0) | -0.1 (-0.1, 0.0) | -0.1 (-0.1, 0.0) | -0.1 (-0.2, 0.0) | -0.1 (-0.2, 0.0) | -0.1 (-0.1, 0.0) | -0.1 (-0.1, 0.0) | -0.1 (-0.1, 0.0) | -0.1 (-0.1, 0.0) | -0.1 (-0.1, 0.0) | -0.1 (-0.1, 0.0) |
| MiAge (1st) | -0.1 (-0.3, 0.0) | -0.1 (-0.3, 0.0) | -0.1 (-0.3, 0.0) | -0.1 (-0.3, 0.0) | -0.1 (-0.3, 0.0) | -0.1 (-0.3, 0.0) | -0.1 (-0.3, 0.0) | -0.1 (-0.3, 0.0) | -0.1 (-0.3, 0.0) | -0.1 (-0.3, 0.0) | -0.1 (-0.3, 0.0) | -0.1 (-0.3, 0.0) | -0.1 (-0.3, 0.0) | -0.1 (-0.3, 0.0) |
| DNAmTL (1st) | 0.3 (0.1, 0.4) | 0.3 (0.1, 0.4) | 0.2 (0.1, 0.4) | 0.2 (0.1, 0.4) | 0.3 (0.1, 0.4) | 0.2 (0.1, 0.4) | 0.2 (0.1, 0.4) | 0.2 (0.1, 0.4) | 0.3 (0.1, 0.4) | 0.3 (0.1, 0.4) | 0.2 (0.1, 0.4) | 0.2 (0.1, 0.4) | 0.3 (0.1, 0.4) | 0.3 (0.1, 0.4) |
| Zhang Mortality (2nd) | 1.6 (1.4, 1.8) | 1.6 (1.4, 1.7) | 1.6 (1.4, 1.8) | 1.6 (1.4, 1.7) | 1.6 (1.4, 1.7) | 1.6 (1.4, 1.7) | 1.6 (1.4, 1.7) | 1.6 (1.4, 1.7) | 1.6 (1.4, 1.7) | 1.6 (1.4, 1.7) | 1.6 (1.4, 1.7) | 1.6 (1.4, 1.8) | 1.6 (1.4, 1.8) | 1.6 (1.4, 1.8) |
| PhenoAge (2nd) | 0.1 (0.0, 0.3) | 0.1 (0.0, 0.3) | 0.1 (0.0, 0.3) | 0.1 (0.0, 0.3) | 0.1 (0.0, 0.3) | 0.1 (0.0, 0.3) | 0.1 (0.0, 0.3) | 0.1 (0.0, 0.3) | 0.1 (0.0, 0.3) | 0.1 (0.0, 0.3) | 0.1 (0.0, 0.3) | 0.1 (0.0, 0.3) | 0.1 (0.0, 0.3) | 0.1 (0.0, 0.3) |
| DunedinPoAm (2nd) | 1.7 (1.5, 1.9) | 1.7 (1.5, 1.9) | 1.7 (1.5, 1.9) | 1.7 (1.5, 1.8) | 1.7 (1.5, 1.9) | 1.7 (1.5, 1.8) | 1.7 (1.5, 1.8) | 1.7 (1.5, 1.9) | 1.7 (1.5, 1.9) | 1.7 (1.5, 1.9) | 1.7 (1.5, 1.8) | 1.7 (1.5, 1.8) | 1.7 (1.5, 1.9) | 1.7 (1.5, 1.8) |
| GrimAge (2nd) | 1.0 (0.9, 1.0) | 1.0 (0.9, 1.0) | 1.0 (0.9, 1.0) | 1.0 (0.9, 1.0) | 1.0 (0.9, 1.0) | 0.9 (0.9, 1.0) | 1.0 (0.9, 1.0) | 1.0 (0.9, 1.0) | 1.0 (0.9, 1.0) | 1.0 (0.9, 1.1) | 1.0 (0.9, 1.0) | 1.0 (0.9, 1.0) | 1.0 (0.9, 1.0) | 1.0 (0.9, 1.0) |

eTable 5 (Part C). Standardized estimates of epigenetic age acceleration associated with age, MBMS, Estimate and 95% CIs
| Clock | Born in a Jim Crow State | Reversed ICE for Racialized Segregation at City of Birth (Black vs. White) | State Policy Conservatism in State of Birth | Parent's Highest Education | Participant's Highest Education | Experiences of Discrimination | Negative log10(household income / poverty line) in 2010 Dollars | Occupational Class | Housing Tenure — Paying Mortgage | Black Carbon | Pollution Proximity Index | Reversed Residential Census Tract ICE for Income | Reversed Residential Census Tract ICE for Racialized Economic Segregation | Reversed Residential Census Tract ICE for Racial Segregation | Reversed Residential Census Tract ICE for Housing Tenure |
| --- | --- | --- | --- | --- | --- | --- | --- | --- | --- | --- | --- | --- | --- | --- | --- |
| Horvath (1st) | 0.6 (0.5, 0.7) | 0.6 (0.5, 0.7) | 0.6 (0.5, 0.7) | 0.6 (0.5, 0.7) | 0.6 (0.5, 0.7) | 0.6 (0.5, 0.7) | 0.6 (0.5, 0.7) | 0.6 (0.5, 0.7) | 0.6 (0.5, 0.7) | 0.6 (0.5, 0.7) | 0.6 (0.5, 0.7) | 0.6 (0.5, 0.7) | 0.6 (0.5, 0.7) | 0.6 (0.5, 0.7) | 0.6 (0.5, 0.7) |
| Hannum (1st) | 0.6 (0.5, 0.7) | 0.6 (0.6, 0.7) | 0.6 (0.5, 0.7) | 0.7 (0.6, 0.8) | 0.6 (0.6, 0.7) | 0.6 (0.6, 0.7) | 0.6 (0.6, 0.7) | 0.6 (0.6, 0.7) | 0.6 (0.6, 0.7) | 0.6 (0.6, 0.7) | 0.7 (0.6, 0.7) | 0.7 (0.6, 0.7) | 0.6 (0.6, 0.7) | 0.6 (0.6, 0.7) | 0.7 (0.6, 0.7) |
| epiToc (1st) | 0.1 (0.0, 0.2) | 0.1 (0.1, 0.2) | 0.1 (0.0, 0.2) | 0.1 (0.1, 0.2) | 0.1 (0.1, 0.2) | 0.1 (0.1, 0.2) | 0.1 (0.1, 0.2) | 0.1 (0.1, 0.2) | 0.1 (0.1, 0.2) | 0.1 (0.1, 0.2) | 0.1 (0.1, 0.2) | 0.1 (0.1, 0.2) | 0.1 (0.1, 0.2) | 0.1 (0.1, 0.2) | 0.1 (0.1, 0.2) |
| Zhang Age (1st) | 0.7 (0.7, 0.8) | 0.8 (0.7, 0.8) | 0.7 (0.7, 0.8) | 0.8 (0.7, 0.8) | 0.8 (0.7, 0.8) | 0.8 (0.7, 0.8) | 0.8 (0.7, 0.8) | 0.8 (0.7, 0.8) | 0.8 (0.7, 0.8) | 0.8 (0.7, 0.8) | 0.8 (0.7, 0.8) | 0.8 (0.7, 0.8) | 0.8 (0.7, 0.8) | 0.8 (0.7, 0.8) | 0.8 (0.7, 0.8) |
| MiAge (1st) | 0.1 (0.0, 0.2) | 0.1 (0.1, 0.2) | 0.1 (0.0, 0.2) | 0.1 (0.0, 0.2) | 0.1 (0.1, 0.2) | 0.1 (0.1, 0.2) | 0.1 (0.1, 0.2) | 0.1 (0.1, 0.2) | 0.1 (0.1, 0.2) | 0.1 (0.1, 0.2) | 0.1 (0.1, 0.2) | 0.1 (0.1, 0.2) | 0.1 (0.1, 0.2) | 0.1 (0.1, 0.2) | 0.1 (0.1, 0.2) |
| DNAmTL (1st) | 0.3 (0.2, 0.4) | 0.4 (0.3, 0.5) | 0.3 (0.3, 0.4) | 0.4 (0.3, 0.5) | 0.4 (0.3, 0.4) | 0.4 (0.3, 0.5) | 0.4 (0.3, 0.5) | 0.4 (0.3, 0.4) | 0.4 (0.3, 0.4) | 0.4 (0.3, 0.5) | 0.4 (0.3, 0.4) | 0.4 (0.3, 0.4) | 0.4 (0.3, 0.4) | 0.4 (0.3, 0.4) | 0.4 (0.3, 0.5) |
| Zhang Mortality (2nd) | 0.0 (-0.1, 0.1) | 0.0 (-0.1, 0.1) | 0.0 (-0.1, 0.1) | 0.1 (0.0, 0.1) | 0.0 (-0.1, 0.1) | 0.0 (-0.1, 0.1) | 0.0 (0.0, 0.1) | 0.0 (0.0, 0.1) | 0.0 (-0.1, 0.1) | 0.0 (-0.1, 0.1) | 0.0 (-0.1, 0.1) | 0.0 (-0.1, 0.1) | 0.0 (-0.1, 0.1) | 0.0 (-0.1, 0.1) | 0.0 (-0.1, 0.1) |
| PhenoAge (2nd) | 0.6 (0.5, 0.6) | 0.6 (0.5, 0.7) | 0.6 (0.5, 0.7) | 0.6 (0.5, 0.7) | 0.6 (0.5, 0.7) | 0.6 (0.5, 0.7) | 0.6 (0.5, 0.7) | 0.6 (0.5, 0.7) | 0.6 (0.5, 0.7) | 0.6 (0.5, 0.7) | 0.6 (0.5, 0.7) | 0.6 (0.5, 0.7) | 0.6 (0.5, 0.7) | 0.6 (0.5, 0.7) | 0.6 (0.5, 0.7) |
| DunedinPoAm (2nd) | -0.1 (-0.2, 0.0) | 0.0 (-0.1, 0.1) | -0.1 (-0.2, 0.0) | 0.0 (-0.1, 0.1) | 0.0 (-0.1, 0.1) | 0.0 (-0.1, 0.1) | 0.0 (-0.1, 0.1) | 0.0 (-0.1, 0.1) | 0.0 (-0.1, 0.1) | 0.0 (-0.1, 0.1) | 0.0 (-0.1, 0.1) | 0.0 (-0.1, 0.1) | 0.0 (-0.1, 0.1) | 0.0 (-0.1, 0.1) | 0.0 (-0.1, 0.1) |
| GrimAge (2nd) | 0.7 (0.7, 0.8) | 0.7 (0.7, 0.8) | 0.7 (0.7, 0.8) | 0.7 (0.7, 0.8) | 0.7 (0.7, 0.8) | 0.7 (0.7, 0.8) | 0.7 (0.7, 0.8) | 0.7 (0.7, 0.8) | 0.7 (0.7, 0.8) | 0.7 (0.7, 0.8) | 0.7 (0.7, 0.8) | 0.7 (0.7, 0.8) | 0.7 (0.7, 0.8) | 0.7 (0.7, 0.8) | 0.7 (0.7, 0.8) |

| Clock | Born in a Jim Crow State | State Policy Conservatism in State of Birth | Parent's Highest Education | Participant's Highest Education | Major Discrimination Scale | Negative log10(household income / poverty line) in 2010 Dollars | Occupational Class | Housing Tenure — Paying Mortgage | Light Absorption Coefficient | Nitrous Oxides | Reversed Residential Census Tract ICE for Income | Reversed Residential Census Tract ICE for Racialized Economic Segregation | Reversed Residential Census Tract ICE for Racial Segregation | Reversed Residential Census Tract ICE for Housing Tenure |
| --- | --- | --- | --- | --- | --- | --- | --- | --- | --- | --- | --- | --- | --- | --- |
| Horvath (1st) | 0.5 (0.5, 0.6) | 0.6 (0.5, 0.6) | 0.6 (0.5, 0.6) | 0.5 (0.5, 0.6) | 0.5 (0.5, 0.6) | 0.6 (0.5, 0.6) | 0.5 (0.5, 0.6) | 0.6 (0.5, 0.6) | 0.5 (0.5, 0.6) | 0.5 (0.5, 0.6) | 0.5 (0.5, 0.6) | 0.5 (0.5, 0.6) | 0.5 (0.5, 0.6) | 0.5 (0.5, 0.6) |
| Hannum (1st) | 0.6 (0.6, 0.7) | 0.6 (0.6, 0.7) | 0.6 (0.6, 0.7) | 0.6 (0.6, 0.7) | 0.6 (0.6, 0.7) | 0.7 (0.6, 0.7) | 0.6 (0.6, 0.7) | 0.7 (0.6, 0.7) | 0.6 (0.6, 0.7) | 0.6 (0.6, 0.7) | 0.6 (0.6, 0.7) | 0.6 (0.6, 0.7) | 0.6 (0.6, 0.7) | 0.6 (0.6, 0.7) |
| epiToc (1st) | 0.1 (0.1, 0.2) | 0.1 (0.1, 0.2) | 0.2 (0.1, 0.2) | 0.1 (0.1, 0.2) | 0.1 (0.1, 0.2) | 0.2 (0.1, 0.2) | 0.2 (0.1, 0.2) | 0.1 (0.1, 0.2) | 0.1 (0.1, 0.2) | 0.1 (0.1, 0.2) | 0.1 (0.1, 0.2) | 0.2 (0.1, 0.2) | 0.2 (0.1, 0.2) | 0.1 (0.1, 0.2) |
| Zhang Age (1st) | 0.8 (0.8, 0.8) | 0.8 (0.8, 0.8) | 0.8 (0.8, 0.8) | 0.8 (0.8, 0.8) | 0.8 (0.8, 0.8) | 0.8 (0.8, 0.8) | 0.8 (0.7, 0.8) | 0.8 (0.8, 0.8) | 0.8 (0.8, 0.8) | 0.8 (0.8, 0.8) | 0.8 (0.8, 0.8) | 0.8 (0.8, 0.8) | 0.8 (0.7, 0.8) | 0.8 (0.8, 0.8) |
| MiAge (1st) | 0.1 (0.1, 0.2) | 0.1 (0.1, 0.2) | 0.1 (0.1, 0.2) | 0.1 (0.1, 0.2) | 0.1 (0.1, 0.2) | 0.1 (0.1, 0.2) | 0.1 (0.1, 0.2) | 0.1 (0.1, 0.2) | 0.1 (0.1, 0.2) | 0.1 (0.1, 0.2) | 0.1 (0.1, 0.2) | 0.1 (0.1, 0.2) | 0.1 (0.1, 0.2) | 0.1 (0.1, 0.2) |
| DNAmTL (1st) | 0.3 (0.3, 0.4) | 0.4 (0.3, 0.4) | 0.3 (0.3, 0.4) | 0.3 (0.3, 0.4) | 0.3 (0.3, 0.4) | 0.3 (0.3, 0.4) | 0.3 (0.3, 0.4) | 0.4 (0.3, 0.4) | 0.4 (0.3, 0.4) | 0.3 (0.3, 0.4) | 0.4 (0.3, 0.4) | 0.4 (0.3, 0.4) | 0.3 (0.3, 0.4) | 0.3 (0.3, 0.4) |
| Zhang Mortality (2nd) | 0.0 (0.0, 0.1) | 0.1 (0.0, 0.1) | 0.0 (0.0, 0.1) | 0.0 (0.0, 0.1) | 0.0 (0.0, 0.1) | 0.0 (0.0, 0.1) | 0.0 (0.0, 0.1) | 0.0 (0.0, 0.1) | 0.0 (0.0, 0.1) | 0.0 (0.0, 0.1) | 0.0 (0.0, 0.1) | 0.0 (0.0, 0.1) | 0.0 (0.0, 0.1) | 0.0 (0.0, 0.1) |
| PhenoAge (2nd) | 0.6 (0.6, 0.7) | 0.6 (0.6, 0.7) | 0.6 (0.6, 0.7) | 0.6 (0.6, 0.7) | 0.6 (0.6, 0.7) | 0.6 (0.6, 0.7) | 0.6 (0.6, 0.7) | 0.6 (0.5, 0.7) | 0.6 (0.6, 0.7) | 0.6 (0.6, 0.7) | 0.6 (0.6, 0.7) | 0.6 (0.6, 0.7) | 0.6 (0.5, 0.6) | 0.6 (0.6, 0.7) |
| DunedinPoAm (2nd) | -0.1 (-0.1, 0.0) | -0.1 (-0.1, 0.0) | -0.1 (-0.1, 0.0) | -0.1 (-0.1, 0.0) | -0.1 (-0.1, 0.0) | -0.1 (-0.1, 0.0) | -0.1 (-0.1, 0.0) | 0.0 (-0.1, 0.0) | -0.1 (-0.1, 0.0) | -0.1 (-0.1, 0.0) | -0.1 (-0.1, 0.0) | -0.1 (-0.1, 0.0) | -0.1 (-0.1, 0.0) | -0.1 (-0.1, 0.0) |
| GrimAge (2nd) | 0.8 (0.7, 0.8) | 0.8 (0.7, 0.8) | 0.8 (0.7, 0.8) | 0.8 (0.7, 0.8) | 0.8 (0.7, 0.8) | 0.7 (0.7, 0.8) | 0.7 (0.7, 0.8) | 0.8 (0.7, 0.8) | 0.8 (0.7, 0.8) | 0.7 (0.7, 0.8) | 0.8 (0.7, 0.8) | 0.8 (0.7, 0.8) | 0.7 (0.7, 0.8) | 0.7 (0.7, 0.8) |
| Horvath (1st) | 0.1 (0.0, 0.2) | 0.1 (0.0, 0.2) | 0.1 (0.0, 0.2) | 0.1 (-0.1, 0.2) | 0.1 (0.0, 0.2) | 0.1 (0.0, 0.2) | 0.1 (0.0, 0.2) | 0.1 (0.0, 0.2) | 0.1 (0.0, 0.2) | 0.1 (0.0, 0.2) | 0.1 (0.0, 0.2) | 0.1 (0.0, 0.2) | 0.1 (0.0, 0.2) | 0.1 (0.0, 0.2) | 0.1 (0.0, 0.2) |
| Hannum (1st) | 0.0 (-0.1, 0.1) | 0.0 (-0.1, 0.1) | 0.0 (-0.1, 0.1) | 0.1 (0.0, 0.2) | 0.0 (-0.1, 0.1) | 0.0 (-0.1, 0.1) | 0.0 (-0.1, 0.1) | 0.0 (-0.1, 0.1) | 0.0 (-0.1, 0.1) | 0.0 (-0.1, 0.1) | 0.0 (-0.1, 0.1) | 0.0 (-0.1, 0.1) | 0.0 (-0.1, 0.1) | 0.0 (-0.1, 0.1) | 0.0 (-0.1, 0.1) |
| epiToc (1st) | 0.1 (0.0, 0.1) | 0.1 (0.0, 0.1) | 0.1 (0.0, 0.1) | 0.1 (0.0, 0.2) | 0.1 (0.0, 0.1) | 0.1 (0.0, 0.1) | 0.1 (0.0, 0.1) | 0.1 (0.0, 0.1) | 0.1 (0.0, 0.1) | 0.1 (0.0, 0.1) | 0.1 (0.0, 0.1) | 0.1 (0.0, 0.1) | 0.1 (0.0, 0.1) | 0.1 (0.0, 0.1) | 0.1 (0.0, 0.1) |
| Zhang Age (1st) | 0.0 (-0.1, 0.0) | 0.0 (-0.1, 0.0) | 0.0 (-0.1, 0.0) | 0.0 (-0.1, 0.1) | 0.0 (-0.1, 0.0) | 0.0 (-0.1, 0.0) | 0.0 (-0.1, 0.0) | 0.0 (-0.1, 0.1) | 0.0 (-0.1, 0.0) | 0.0 (-0.1, 0.0) | 0.0 (-0.1, 0.0) | 0.0 (-0.1, 0.1) | 0.0 (-0.1, 0.1) | 0.0 (-0.1, 0.0) | 0.0 (-0.1, 0.1) |
| MiAge (1st) | 0.1 (0.0, 0.1) | 0.1 (0.0, 0.1) | 0.1 (0.0, 0.1) | 0.0 (-0.1, 0.1) | 0.1 (0.0, 0.1) | 0.1 (0.0, 0.1) | 0.1 (0.0, 0.1) | 0.1 (0.0, 0.1) | 0.1 (0.0, 0.1) | 0.1 (0.0, 0.1) | 0.1 (0.0, 0.1) | 0.1 (0.0, 0.2) | 0.1 (0.0, 0.2) | 0.1 (0.0, 0.1) | 0.1 (0.0, 0.2) |
| DNAmTL (1st) | -0.1 (-0.2, 0.0) | -0.1 (-0.2, 0.0) | -0.1 (-0.2, 0.0) | -0.2 (-0.3, -0.1) | -0.2 (-0.2, -0.1) | -0.1 (-0.2, 0.0) | -0.1 (-0.2, 0.0) | -0.1 (-0.2, 0.0) | -0.1 (-0.2, 0.0) | -0.1 (-0.2, 0.0) | -0.1 (-0.2, 0.0) | -0.1 (-0.2, 0.0) | -0.1 (-0.2, 0.0) | -0.1 (-0.2, 0.0) | -0.1 (-0.2, 0.0) |
| Zhang Mortality (2nd) | 0.0 (-0.1, 0.1) | 0.0 (-0.1, 0.1) | 0.0 (-0.1, 0.1) | 0.0 (-0.1, 0.1) | 0.0 (-0.1, 0.1) | 0.0 (-0.1, 0.1) | 0.0 (-0.1, 0.1) | 0.0 (-0.1, 0.1) | 0.0 (-0.1, 0.1) | 0.0 (-0.1, 0.1) | 0.0 (-0.1, 0.1) | 0.0 (-0.1, 0.1) | 0.0 (-0.1, 0.1) | 0.0 (-0.1, 0.1) | 0.0 (-0.1, 0.1) |
| PhenoAge (2nd) | 0.1 (0.0, 0.1) | 0.1 (0.0, 0.2) | 0.1 (0.0, 0.2) | 0.1 (0.0, 0.2) | 0.1 (0.0, 0.2) | 0.1 (0.0, 0.1) | 0.1 (0.0, 0.2) | 0.1 (0.0, 0.2) | 0.1 (0.0, 0.2) | 0.1 (0.0, 0.2) | 0.1 (0.0, 0.2) | 0.1 (0.0, 0.2) | 0.1 (0.0, 0.2) | 0.1 (0.0, 0.2) | 0.1 (0.0, 0.2) |
| DunedinPoAm (2nd) | 0.0 (-0.1, 0.1) | 0.0 (-0.1, 0.1) | 0.0 (-0.1, 0.1) | 0.0 (-0.1, 0.1) | 0.0 (-0.1, 0.1) | 0.0 (-0.1, 0.1) | 0.0 (-0.1, 0.1) | 0.0 (-0.1, 0.1) | 0.0 (-0.1, 0.1) | 0.0 (-0.1, 0.1) | 0.0 (-0.1, 0.1) | 0.0 (-0.1, 0.1) | 0.0 (-0.1, 0.1) | 0.0 (-0.1, 0.1) | 0.0 (-0.1, 0.1) |
| GrimAge (2nd) | 0.0 (-0.1, 0.1) | 0.0 (-0.1, 0.1) | 0.0 (-0.1, 0.1) | 0.0 (-0.1, 0.1) | 0.0 (-0.1, 0.1) | 0.0 (-0.1, 0.1) | 0.0 (-0.1, 0.1) | 0.0 (-0.1, 0.1) | 0.0 (-0.1, 0.1) | 0.0 (-0.1, 0.1) | 0.0 (-0.1, 0.0) | 0.0 (-0.1, 0.1) | 0.0 (-0.1, 0.1) | 0.0 (-0.1, 0.1) | 0.0 (-0.1, 0.1) |

| Clock | Born in a Jim Crow State | State Policy Conservatism in State of Birth | Parent's Highest Education | Participant's Highest Education | Major Discrimination Scale | Negative log10(household income / poverty line) in 2010 Dollars | Occupational Class | Housing Tenure — Paying Mortgage | Light Absorption Coefficient | Nitrous Oxides | Reversed Residential Census Tract ICE for Income | Reversed Residential Census Tract ICE for Racialized Economic Segregation | Reversed Residential Census Tract ICE for Racial Segregation | Reversed Residential Census Tract ICE for Housing Tenure |
| --- | --- | --- | --- | --- | --- | --- | --- | --- | --- | --- | --- | --- | --- | --- |
| Horvath (1st) | 0.1 (0.0, 0.1) | 0.0 (0.0, 0.1) | 0.1 (0.0, 0.1) | 0.1 (0.0, 0.1) | 0.1 (0.0, 0.1) | 0.1 (0.0, 0.1) | 0.0 (0.0, 0.1) | 0.0 (0.0, 0.1) | 0.0 (0.0, 0.1) | 0.0 (0.0, 0.1) | 0.1 (0.0, 0.1) | 0.1 (0.0, 0.1) | 0.1 (0.0, 0.1) | 0.1 (0.0, 0.1) |
| Hannum (1st) | 0.0 (0.0, 0.0) | 0.0 (0.0, 0.0) | 0.0 (0.0, 0.0) | 0.0 (0.0, 0.0) | 0.0 (0.0, 0.0) | 0.0 (0.0, 0.0) | 0.0 (0.0, 0.0) | 0.0 (0.0, 0.0) | 0.0 (0.0, 0.0) | 0.0 (0.0, 0.0) | 0.0 (0.0, 0.0) | 0.0 (0.0, 0.0) | 0.0 (0.0, 0.0) | 0.0 (0.0, 0.0) |
| epiToc (1st) | 0.0 (-0.1, 0.0) | 0.0 (-0.1, 0.0) | 0.0 (-0.1, 0.0) | 0.0 (-0.1, 0.0) | 0.0 (-0.1, 0.0) | 0.0 (-0.1, 0.0) | 0.0 (-0.1, 0.0) | 0.0 (-0.1, 0.0) | 0.0 (-0.1, 0.0) | 0.0 (-0.1, 0.0) | 0.0 (-0.1, 0.0) | 0.0 (-0.1, 0.0) | 0.0 (-0.1, 0.0) | 0.0 (-0.1, 0.0) |
| Zhang Age (1st) | 0.0 (0.0, 0.0) | 0.0 (0.0, 0.0) | 0.0 (0.0, 0.0) | 0.0 (0.0, 0.0) | 0.0 (0.0, 0.0) | 0.0 (0.0, 0.0) | 0.0 (0.0, 0.0) | 0.0 (0.0, 0.0) | 0.0 (0.0, 0.0) | 0.0 (0.0, 0.0) | 0.0 (0.0, 0.0) | 0.0 (0.0, 0.0) | 0.0 (0.0, 0.0) | 0.0 (0.0, 0.0) |
| MiAge (1st) | 0.0 (-0.1, 0.0) | 0.0 (-0.1, 0.0) | 0.0 (-0.1, 0.0) | 0.0 (-0.1, 0.0) | 0.0 (-0.1, 0.0) | 0.0 (-0.1, 0.0) | 0.0 (-0.1, 0.0) | 0.0 (-0.1, 0.0) | 0.0 (-0.1, 0.0) | 0.0 (-0.1, 0.0) | 0.0 (-0.1, 0.0) | 0.0 (-0.1, 0.0) | 0.0 (-0.1, 0.0) | 0.0 (-0.1, 0.0) |
| DNAmTL (1st) | 0.0 (0.0, 0.1) | 0.0 (0.0, 0.1) | 0.0 (0.0, 0.1) | 0.0 (0.0, 0.1) | 0.0 (0.0, 0.1) | 0.0 (0.0, 0.1) | 0.0 (0.0, 0.1) | 0.0 (0.0, 0.1) | 0.0 (0.0, 0.1) | 0.0 (0.0, 0.1) | 0.0 (0.0, 0.1) | 0.0 (0.0, 0.1) | 0.0 (0.0, 0.1) | 0.0 (0.0, 0.1) |
| Zhang Mortality (2nd) | 0.0 (0.0, 0.1) | 0.0 (0.0, 0.1) | 0.0 (0.0, 0.1) | 0.0 (0.0, 0.1) | 0.0 (0.0, 0.1) | 0.0 (0.0, 0.1) | 0.0 (0.0, 0.1) | 0.0 (0.0, 0.1) | 0.0 (0.0, 0.1) | 0.0 (0.0, 0.1) | 0.0 (0.0, 0.1) | 0.0 (0.0, 0.1) | 0.0 (0.0, 0.1) | 0.0 (0.0, 0.1) |
| PhenoAge (2nd) | 0.1 (0.0, 0.1) | 0.1 (0.0, 0.1) | 0.1 (0.0, 0.1) | 0.1 (0.0, 0.1) | 0.1 (0.0, 0.1) | 0.1 (0.0, 0.1) | 0.1 (0.0, 0.1) | 0.1 (0.0, 0.1) | 0.1 (0.0, 0.1) | 0.1 (0.0, 0.1) | 0.1 (0.0, 0.1) | 0.1 (0.0, 0.1) | 0.1 (0.0, 0.1) | 0.1 (0.0, 0.1) |
| DunedinPoAm (2nd) | 0.1 (0.0, 0.1) | 0.1 (0.0, 0.1) | 0.1 (0.0, 0.1) | 0.1 (0.0, 0.1) | 0.1 (0.0, 0.1) | 0.1 (0.0, 0.1) | 0.1 (0.0, 0.1) | 0.1 (0.0, 0.1) | 0.1 (0.0, 0.1) | 0.1 (0.0, 0.1) | 0.1 (0.0, 0.1) | 0.1 (0.0, 0.1) | 0.1 (0.0, 0.1) | 0.1 (0.0, 0.1) |
| GrimAge (2nd) | 0.0 (0.0, 0.1) | 0.0 (0.0, 0.1) | 0.0 (0.0, 0.1) | 0.0 (0.0, 0.1) | 0.0 (0.0, 0.1) | 0.0 (0.0, 0.1) | 0.0 (0.0, 0.1) | 0.0 (0.0, 0.1) | 0.0 (0.0, 0.1) | 0.0 (0.0, 0.1) | 0.0 (0.0, 0.1) | 0.0 (0.0, 0.1) | 0.0 (0.0, 0.1) | 0.0 (0.0, 0.1) |

**eTextbox 1. Construction of surrogate variables and genetic principal component (PC) variables.**

**Surrogate variables:** We adjusted for batch effects using surrogate variables (SVs) (the first 5 for MBMS and the first 10 for MESA, calculated using the R package sva [1]). We used SVs to adjust for batch because stratifying the dataset meant that measured batch variables had small cell frequencies, which in turn caused uncertainty in the regression model and resulted in deflation of test statistics. We chose to use 5 SVs for MBMS and 10 for MESA as these encompassed the majority of association between SVs and batch variables, as well as DNA input level for MBMS; and their inclusion returned EWAS lambda values to approximately 1.

[1] Leek JT, Johnson WE, Parker HS, Fertig EJ, Jaffe AE, Zhang Y, Storey JD, Torres LC (2023). *sva: Surrogate Variable Analysis*. R package version 3.48.0.

**Genetic principal component (PC) variables:** MESA used the Affymetrix Genome-Wide Human SNP Array 6.0 to genotype 8402 participants; this overlaps with 933 out of 975 of the US-born participants with DNAm data. Genetic principal components were generated by MESA: 23,428 flagged SNPs and 6,849 SNPs in long range LD were removed before principal components were computed per chromosome, and then combined across chromosomes to give the final PC values. We used the genetic PCs provided to us by MESA that were estimated in all participants combined, as all participants were included together in the models.

Of note, we employed the SVs in the primary analyses because these could be used in both the MBMS and MESA analyses; by contrast, the genetic PCs could be employed solely with the MESA data.

**eTextbox 2. Methods for multiple imputation and multiple comparisons.**

**1) Multiple imputation**

Multiple imputation was carried out in all models to account for missing exposures and covariates. The extent of missing data on each of the variables is listed in Table 1. The imputation models included the epigenetic clock values, age, racialized group, each exposure interacted with racialized groups, BMI, smoking, sex/gender, the cell type variables, and the surrogate variables (5 for MBMS and 10 for MESA). All analytic variables were included in the imputation models andiInteractions were included to reduce the potential for bias. The default, predictive mean matching, was used as the imputation method for each missing variable and 40 imputations were carried out per model. Multiple imputations were performed using the mice package in R.

References:

Buuren S, Groothuis-Oudshoorn K. mice: Multivariate Imputation by Chained Equations in R*. J Statistical Software* 2011; 45(3):1-67.

Tilling K, Williamson EJ, Spratt M, Sterne JAC, Carpenter JR. Appropriate inclusion of interactions was needed to avoid bias in multiple imputation. *J Clin Epidemiol* 2016; 80:107–115.

**2) Multiple comparisons**

The number of multiple comparisons varied and depended on four aspects of each set of analyses: (1) the set of variables considered (e.g., for early life or for adult life exposures), (2) which population group was the focus (MBMS or MESA), (3) the number of racialized groups at issue, and (4) whether the analyses focused on the epigenetic clocks individually or the pooled estimates. As a result, depending on the specific analysis at issue, the number of multiple comparisons ranged between 12 and 290. We list the N of multiple comparisons for each set of analyses below; further details (e.g., specific variables included) can be provided upon request.

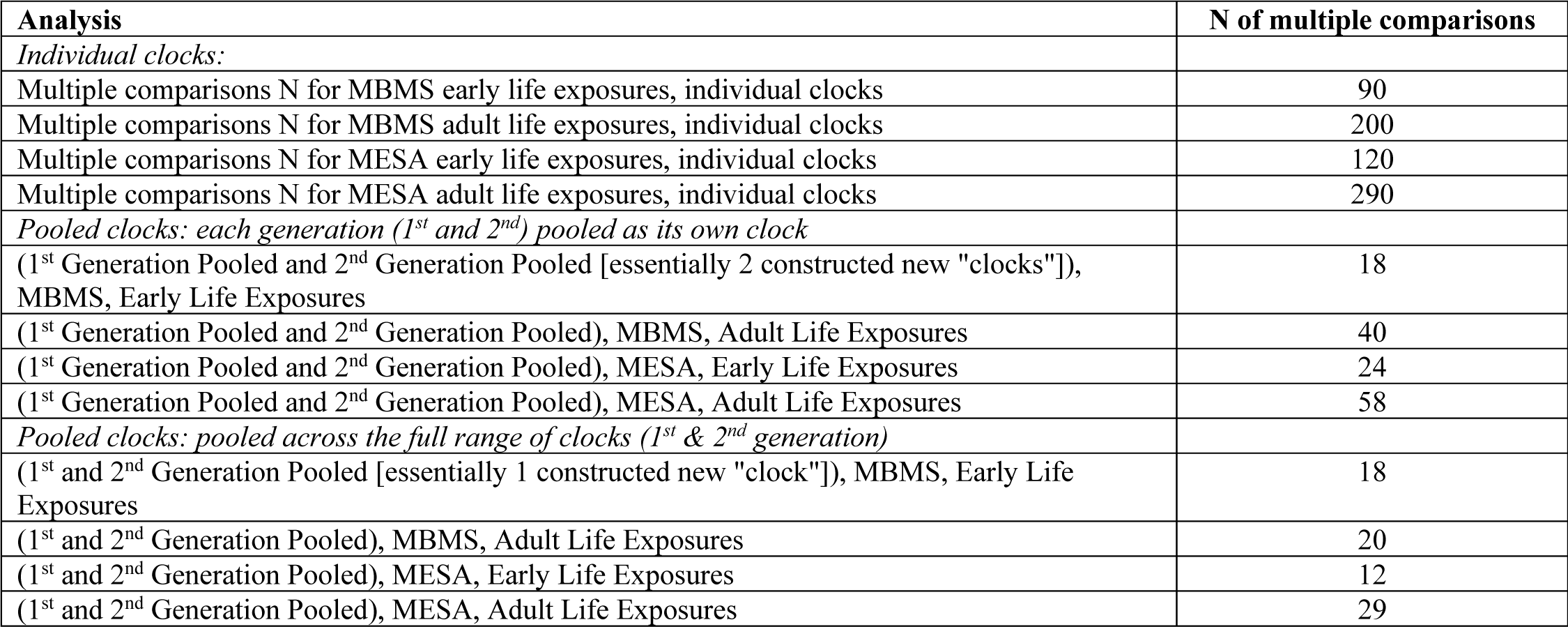

### 2) Supplementary Figures

**eFigure 1.**
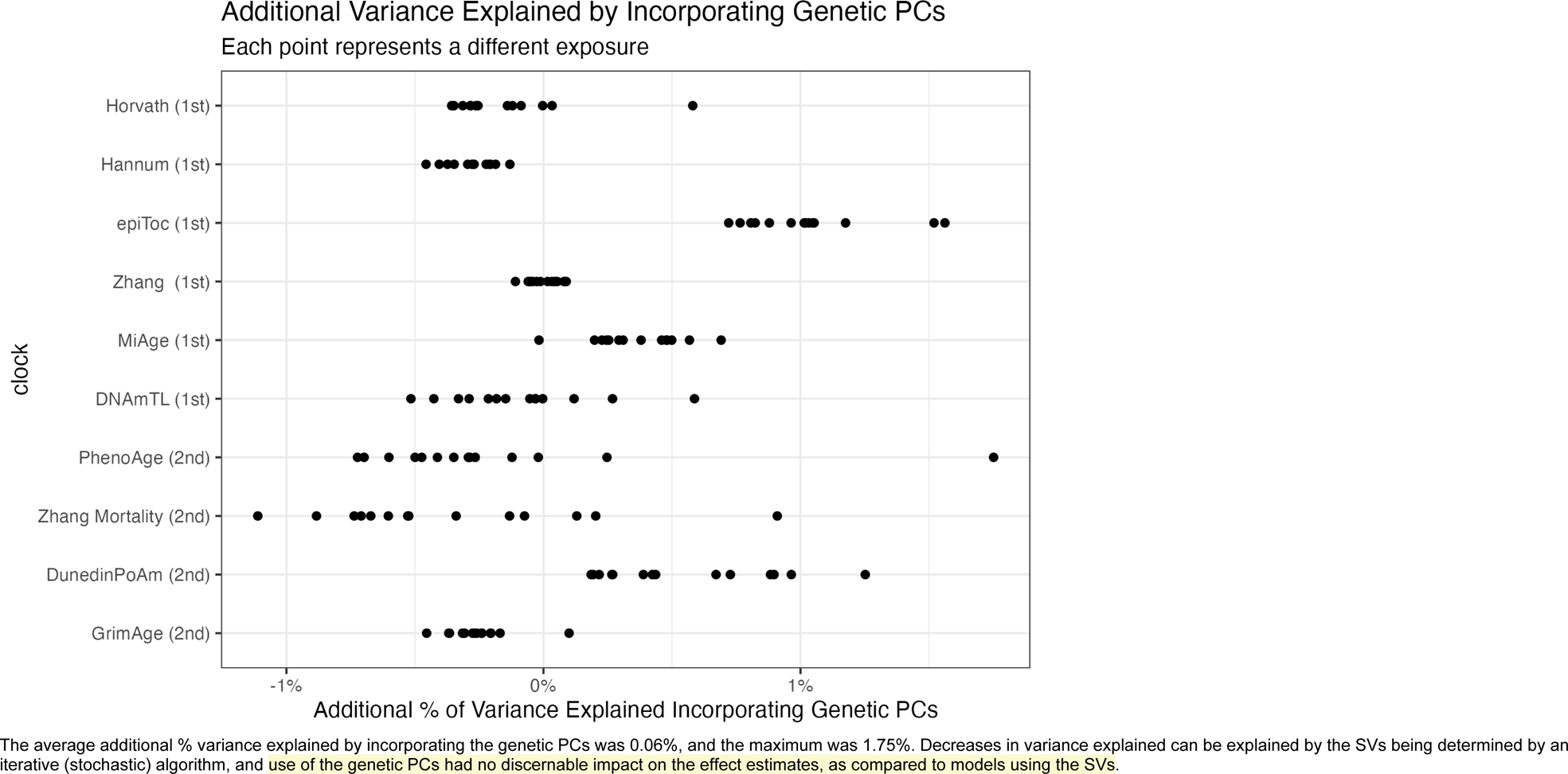
Additional variance explained by incorporating genetic PCs.

**eFigure 2.**
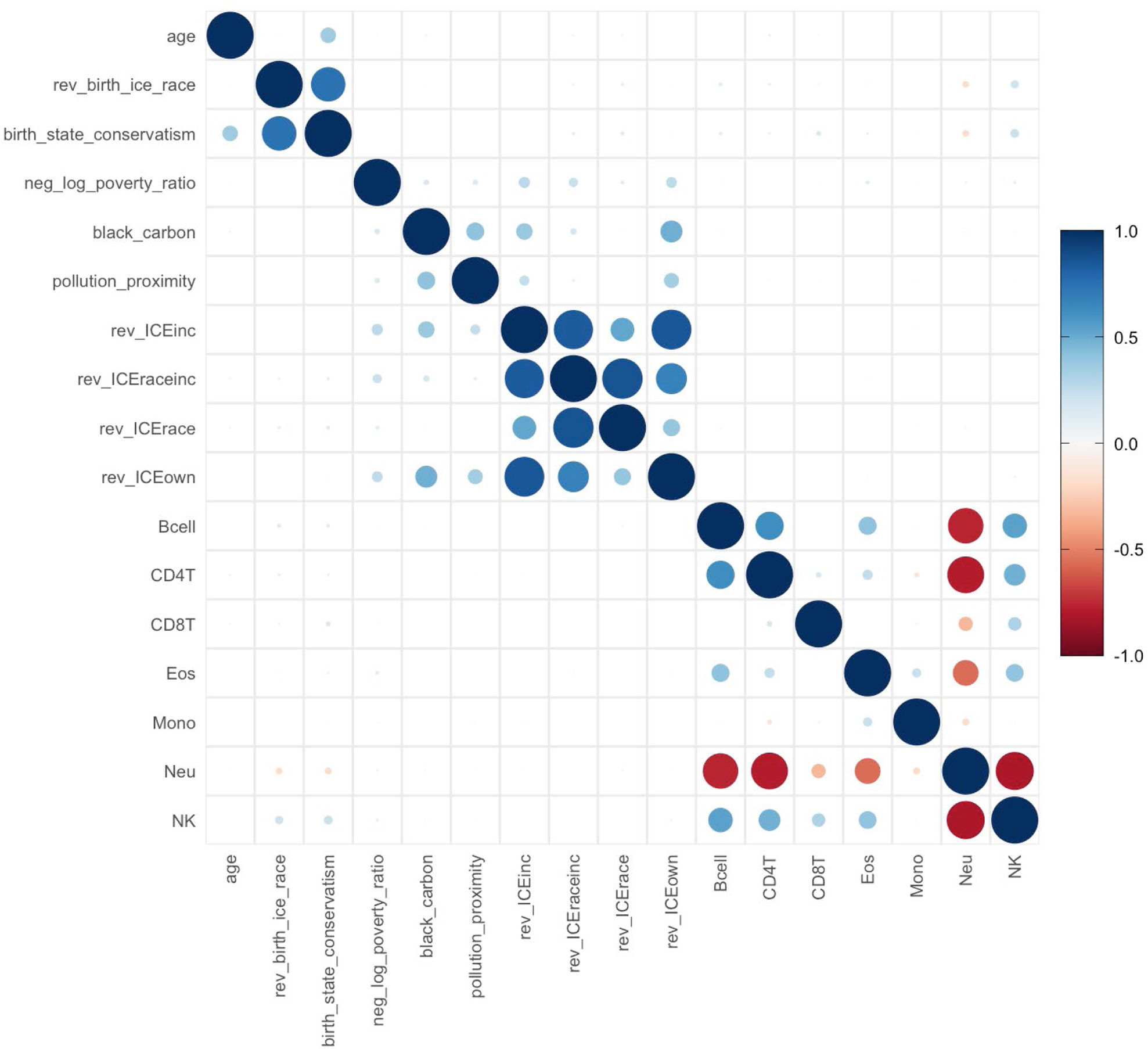
Correlation matrix for continuous variables from MBMS

**eFigure 3.**
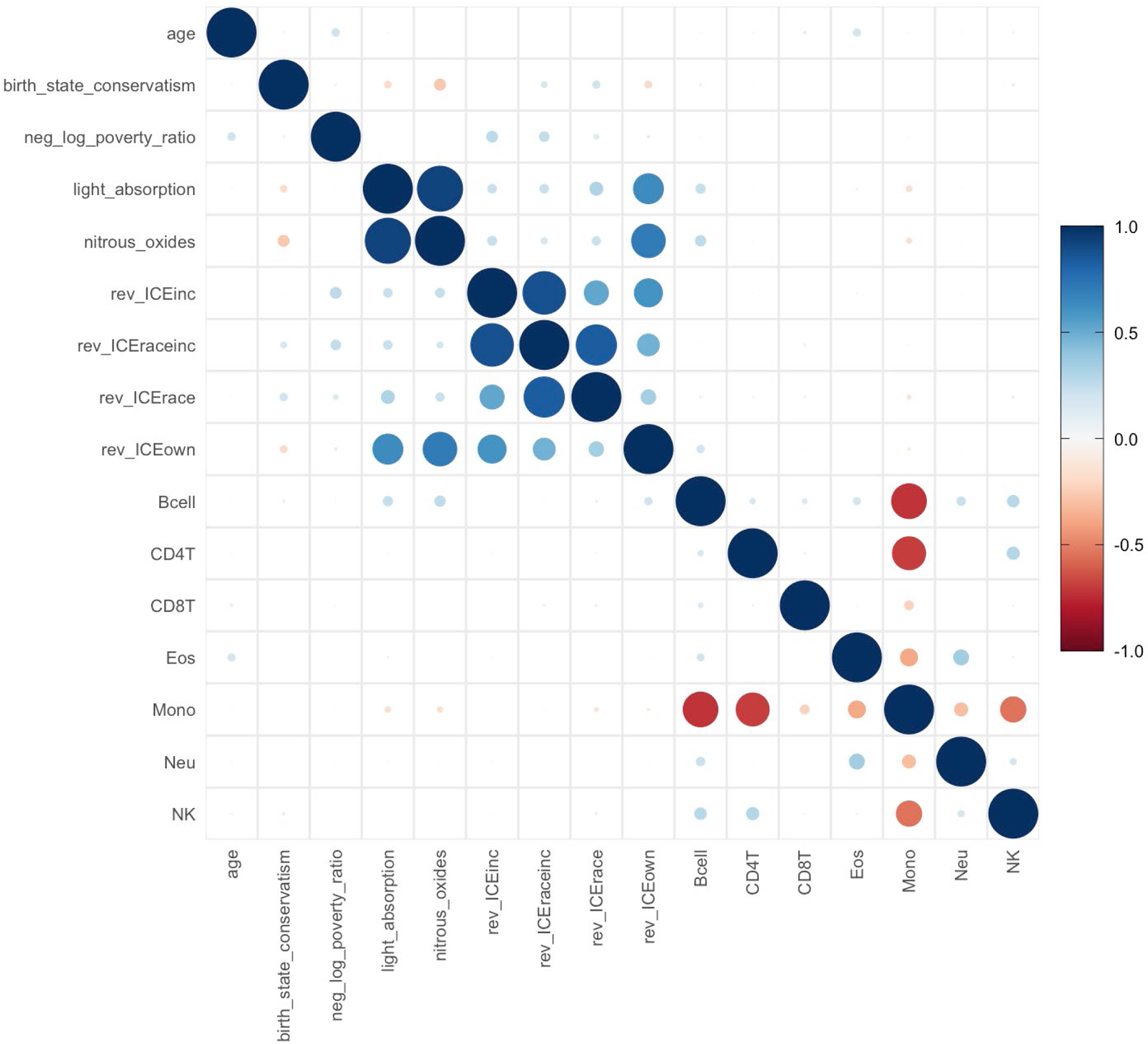
Correlation matrix for continuous variables from MESA, US-Born

**eFigure 4.**
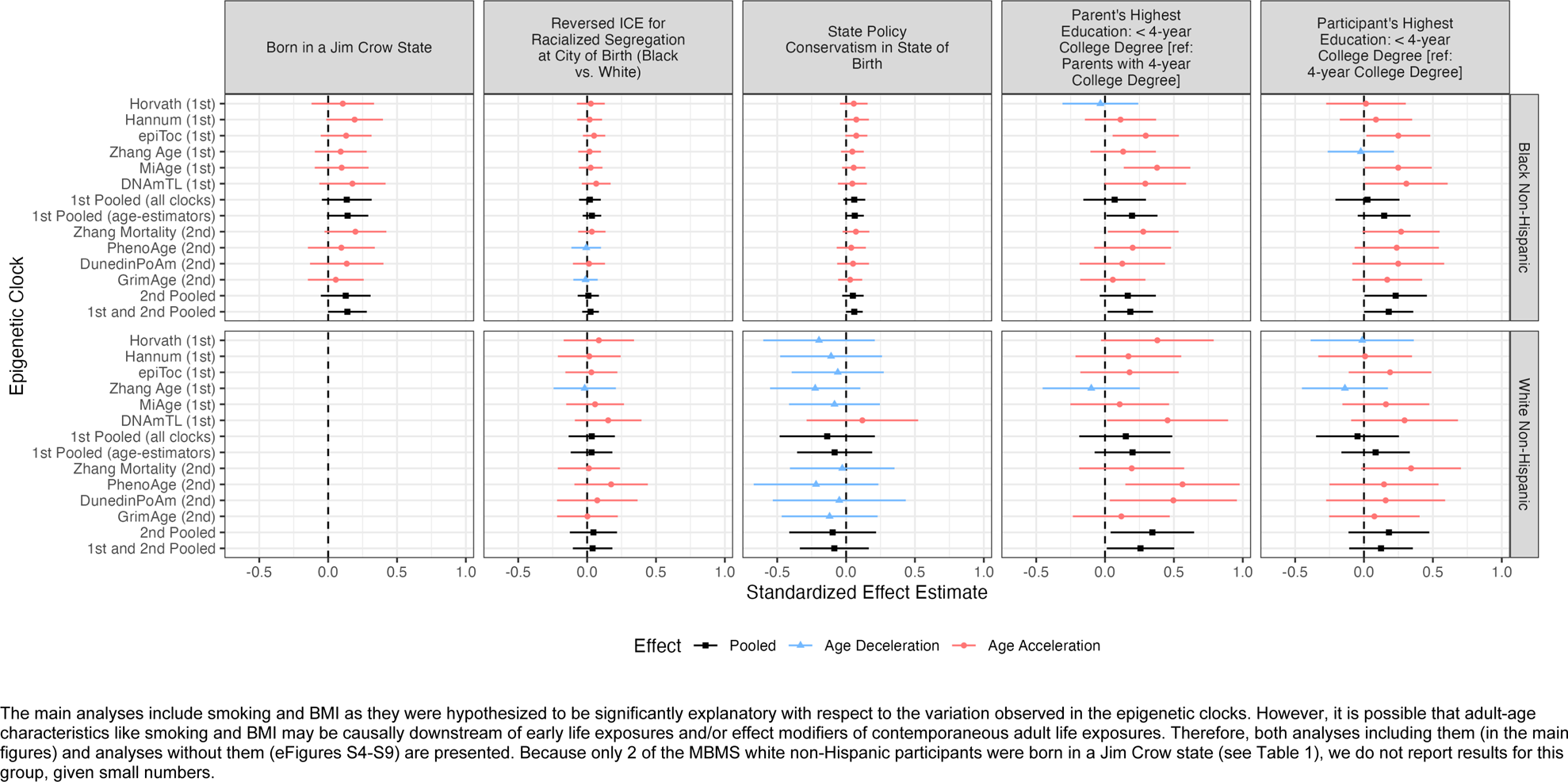
Standardized effect estimates by racialized group and 95% confidence intervals for early life exposures (born in a Jim Crow state, reversed Index of Concentration at the Extremes for racialized segregation at city of birth, state policy conservatism in state of birth, parents’ education, participants’ education): MBMS – not including covariate data on BMI and smoking.

**eFigure 5.**
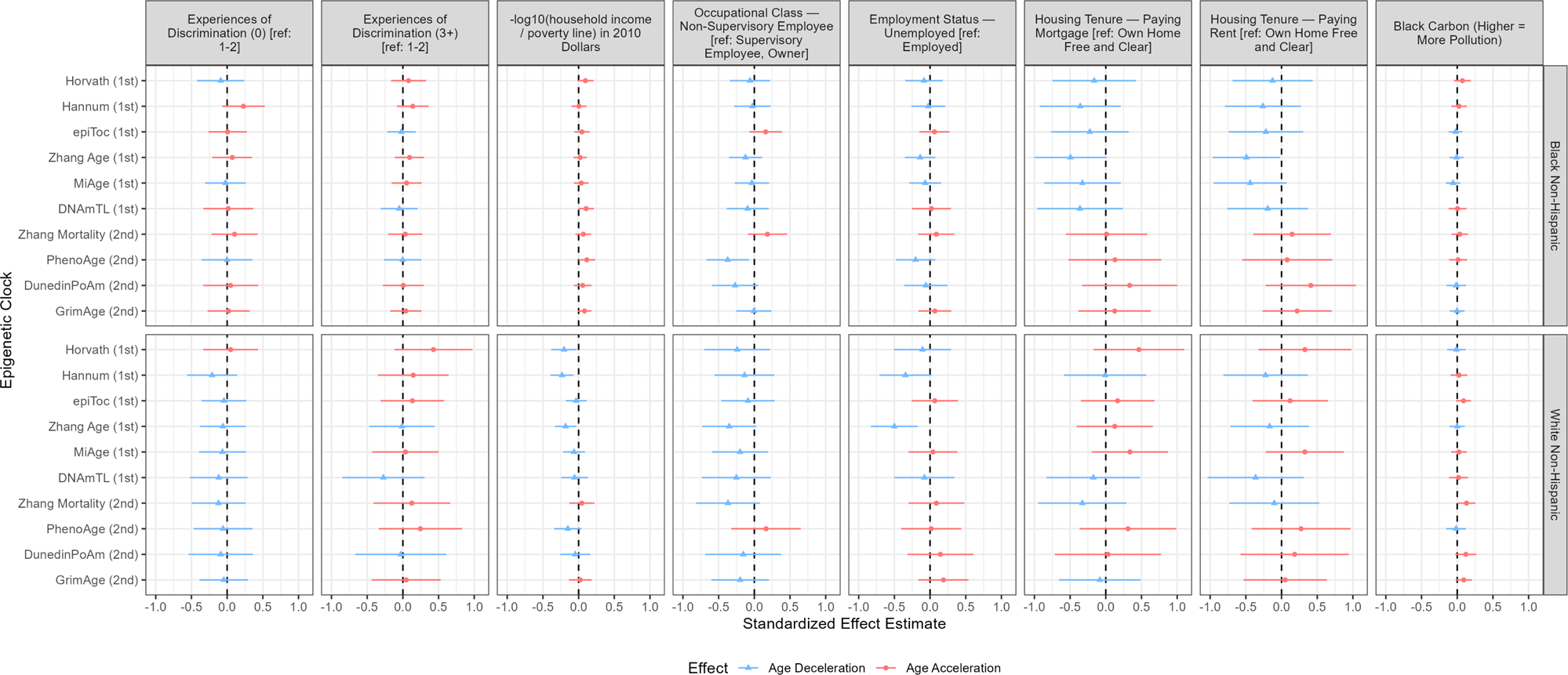
Standardized effect estimates by racialized group and 95% confidence intervals for adult life exposures (set 1: Experiences of Discrimination, negative log household income ratio to the poverty line, occupational class, employment status, housing tenure, and black carbon air pollution): MBMS – not including covariate data on BMI and smoking.

**eFigure 6.**
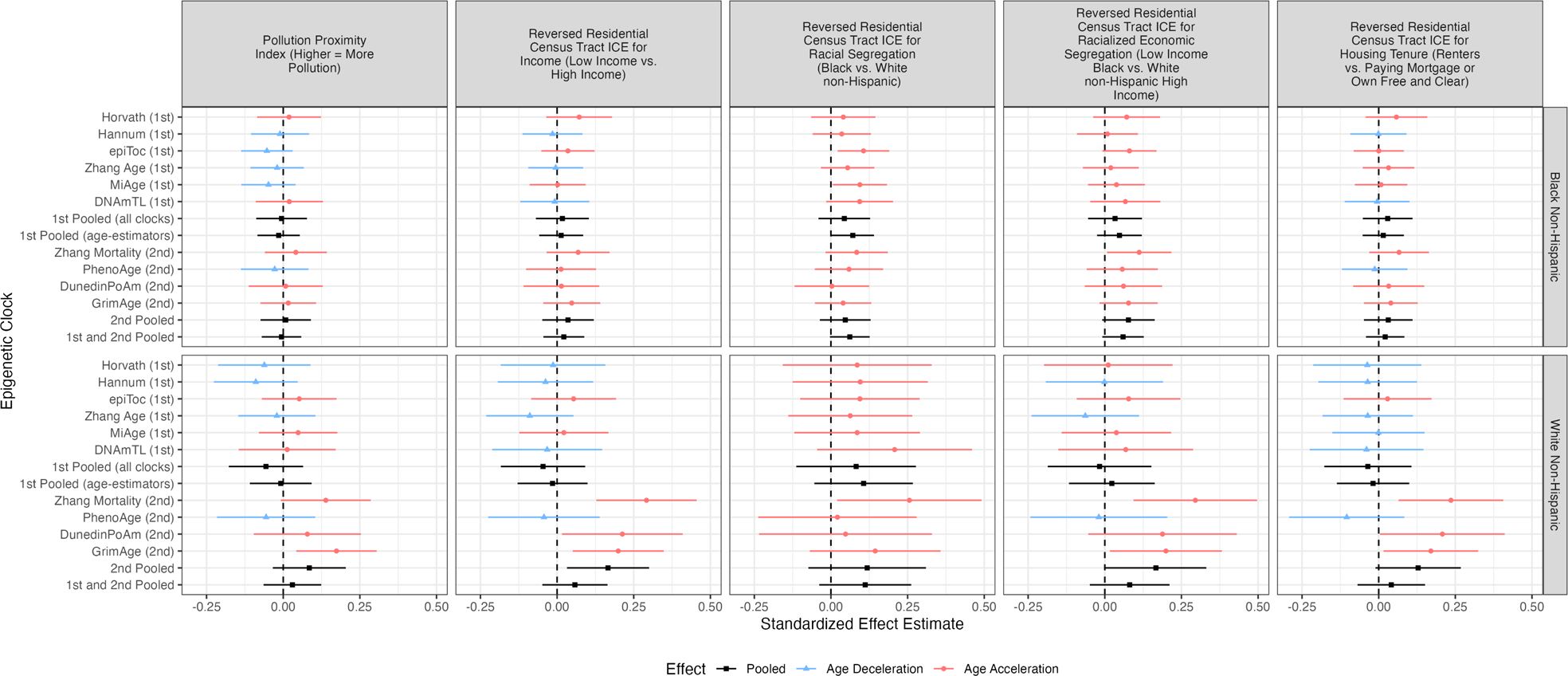
Standardized effect estimates by racialized group and 95% confidence intervals for adult life exposures (set 2: Pollution Proximity Index [air pollution], reversed Index of Concentration at the Extremes [ICE] for: income, racial segregation, racialized economic segregation, and housing tenure): MBMS – not including covariate data on BMI and smoking.

**eFigure 7.**
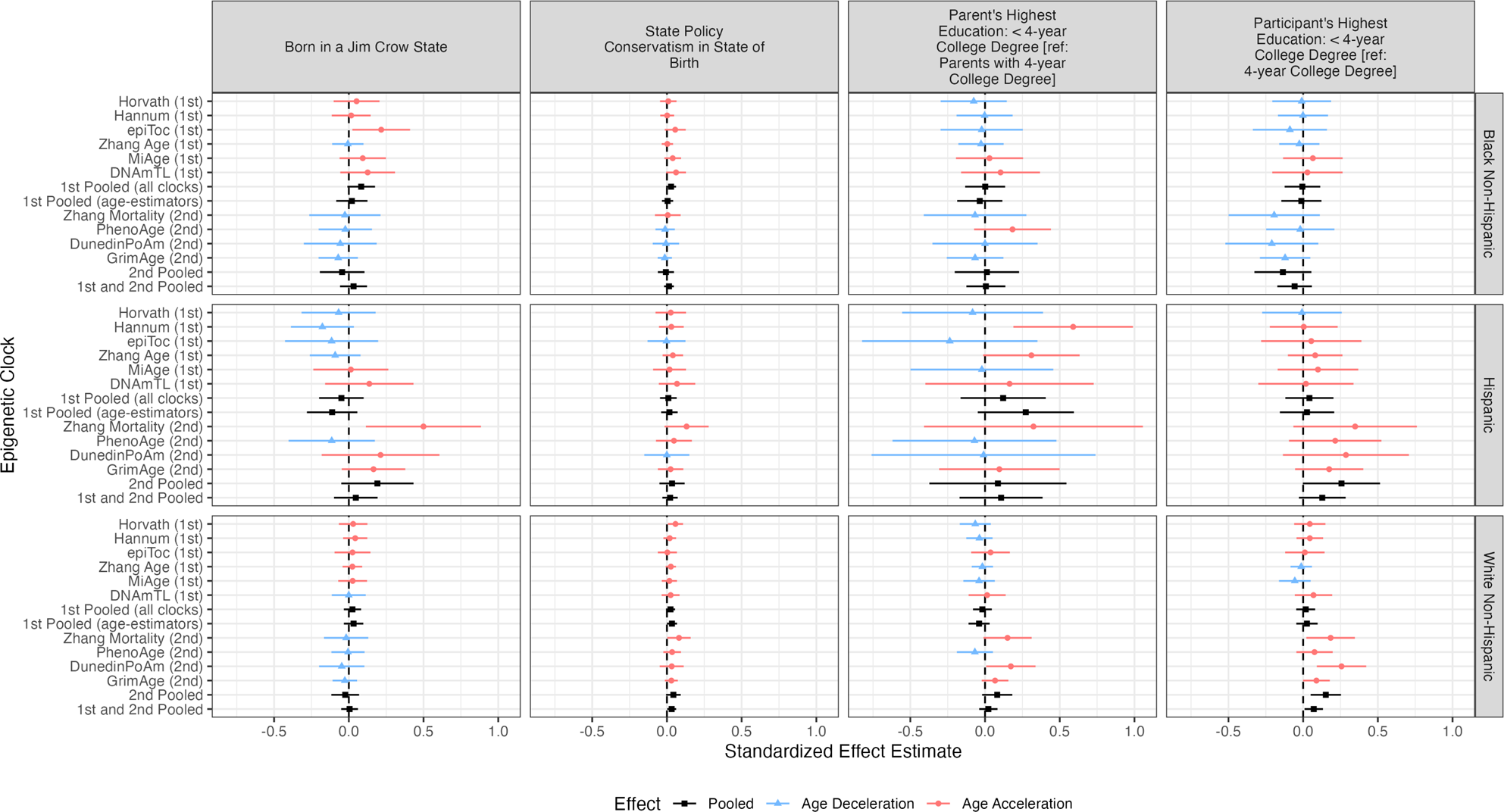
Standardized effect estimates by racialized group and 95% confidence intervals for early life exposures (born in a Jim Crow state, state policy conservatism in state of birth, parents’ education, participants’ education): MESA US-Born – not including covariate data on BMI and smoking.

**eFigure 8.**
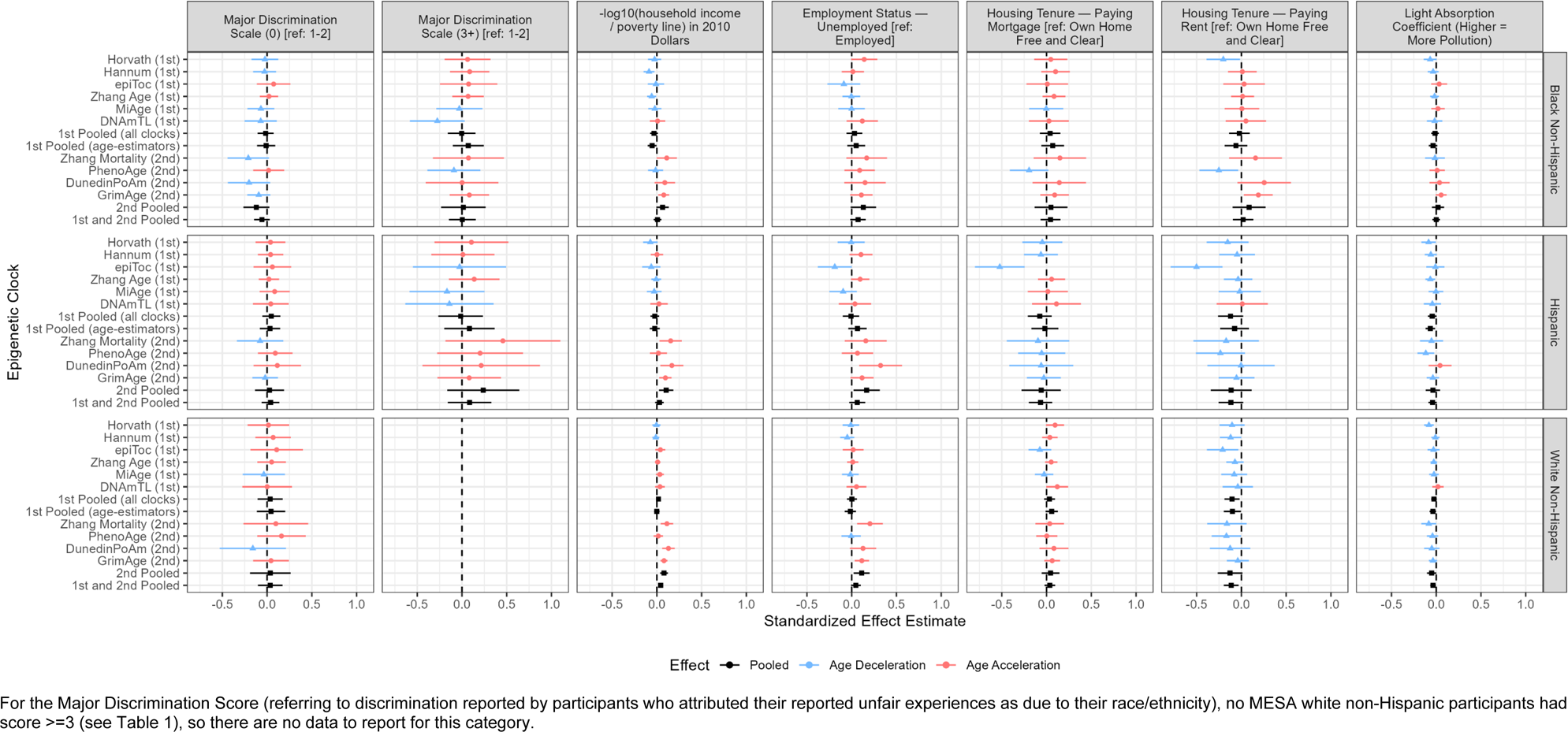
Standardized effect estimates by racialized group and 95% confidence intervals for adult life exposures (set 1: Major Discrimination Scale (racialized), negative log household income ratio to the poverty line, employment status, housing tenure, and light absorption coefficient [air pollution]): MESA US-Born – not including covariate data on BMI and smoking.

**eFigure 9.**
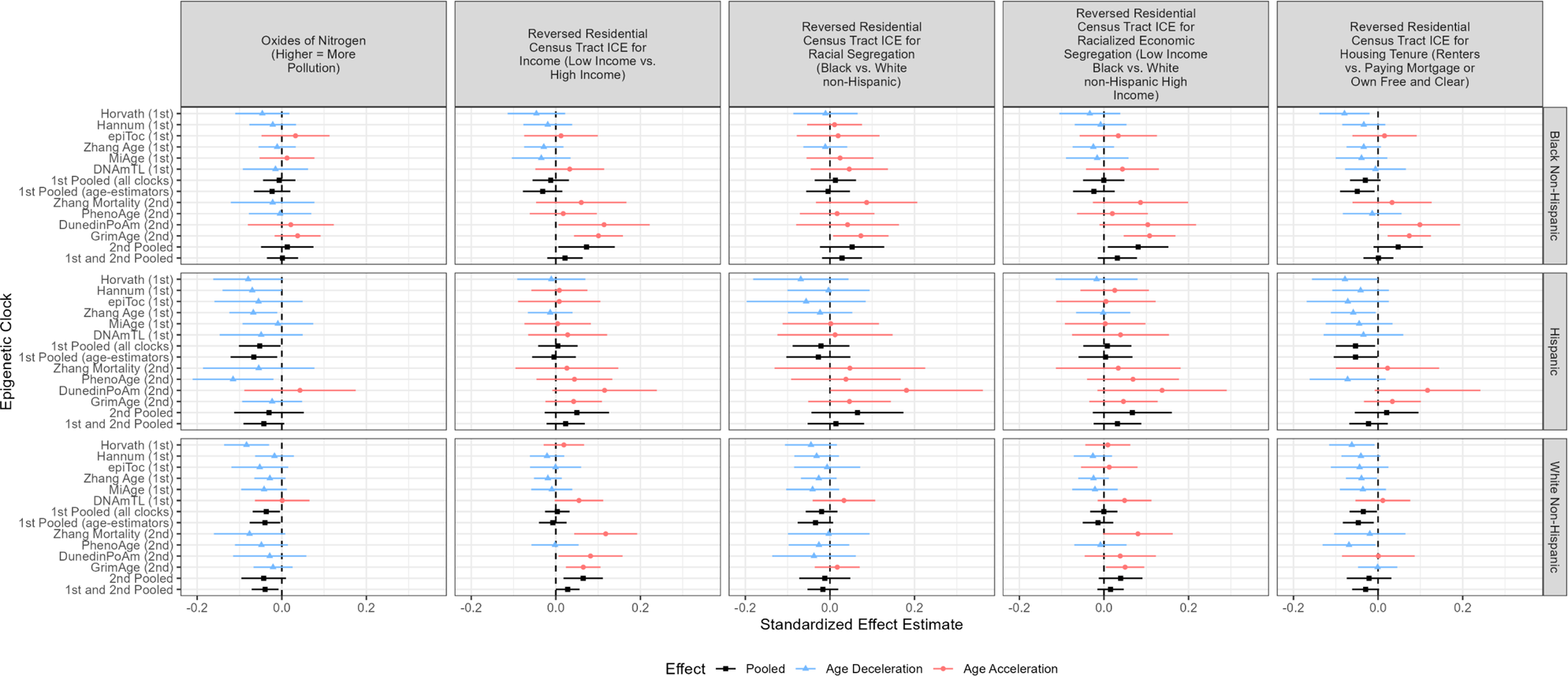
Standardized effect estimates by racialized group and 95% confidence intervals for adult life exposures (set 2: oxides of nitrogen [air pollution], reversed Index of Concentration at the Extremes [ICE] for: income, racial segregation, racialized economic segregation, and housing tenure): MESA US-Born – not including covariate data on BMI and smoking.

## REFERENCES

1. World Health Organization. Health Equity. https://www.who.int/health-topics/health-equity#tab=tab_1; accessed November 28, 2023.

2. Krieger N. Ecosocial Theory, Embodied Truths, and the People’s Health. New York: Oxford University Press, 2021.

3. Relton CL, Hartwig FP, Davey Smith G. From stem cells to the law courts: DNA methylation, the forensic epigenome and the possibility of a biosocial archive. Int J Epidemiol 2015; 44(4):1083–1093.

4. Peixoto P, Cartron PF, Serandour AA, Hervouet E. From 1957 to nowadays: a brief history of epigenetics. Int J Mol Sci 2020; 21(20):7571. doi: 10.3390/ijms21207571.

5. Waddington CH. Organisers & Genes. Cambridge, UK: Cambridge University Press, 1940.

6. Dor Y, Cedar H. Principles of DNA methylation and their implications for biology and medicine. Lancet 2018; 392(10149):777–786.

7. Horvath S, Raj K. DNA methylation-based biomarkers and the epigenetic clock theory of ageing. Nature Review Genetics 2018; 19:371–384.

8. Ryan CP. “Epigenetic clocks”: Theory and applications in human biology. Am J Hum Biol 2021; 33(3):e23488. doi: 10.1002/ajhb.23488.

9. Horvath S. DNA methylation age of human tissues and cell types. Genome Biol 2013; 14:R115.

10. Hannum G, Guinney J, Zhao L, Zhang L, Hughes G. Genome-wide methylation profiles reveal quantitative views of human aging rates. Mol Cell 2013; 49:359–367.

11. Raffington L, Belsky DW. Integrating DNA methylation measures of biological aging into social determinants of health research. Curr Environ Health Rep 2022; 9(2):196–210.

12. Martin CL, Ghastine L, Lodge EK, Dhingra R, Ward-Caviness CK. Understanding health inequalities through the lens of social epigenetics. Annu Rev Public Health 2022; 43:235–254.

13. Oblak L, van der Zaag J, Higgins-Chen AT, Levine ME, Boks MP. A systematic review of biological, social and environmental factors associated with epigenetic clock acceleration. Ageing Res Rev 2021; 69:101348. doi: 10.1016/j.arr.2021.101348

14. Ryan J, Wrigglesworth J, Loong J, Fransquet PD, Woods RL. A systematic review and meta-analysis of environmental, lifestyle, and health factors associated with DNA Methylation Age. J Gerontol A Biol Sci Med Sci 2020; 75(3):481–494.

15. Lo YH, Lin WY. Cardiovascular health and four epigenetic clocks. Clin Epigenetics 2022 Jun 9;14(1):73. doi: 10.1186/s13148-022-01295-7.

16. Faul JD, Kim JK, Levine ME, Thyagarajan B, Weir DR, Crimmins EM. Epigenetic-based age acceleration in a representative sample of older Americans: Associations with aging-related morbidity and mortality. Proc Natl Acad Sci U S A. 2023; 120(9):e2215840120. doi: 10.1073/pnas.2215840120.

17. Lawrence KG, Kresovich JK, O’Brien KM, Hoang TT, Xu Z, Taylor JA, Sandler DP. Association of neighborhood deprivation with epigenetic aging using 4 clock metrics. JAMA Netw Open 2020; 3(11):e2024329. doi: 10.1001/jamanetworkopen.2020.24329

18. White AJ, Kresovich JK, Keller JP, Xu Z, Kaufman JD, Weinberg CR, Taylor JA, Sandler DP. Air pollution, particulate matter composition and methylation-based biologic age. Environ Int 2019; 132:105071. doi: 10.1016/j.envint.2019.105071.

19. Lim S, Nzegwu D, Wright ML. The impact of psychosocial stress from life trauma and racial discrimination on epigenetic aging-a systematic review. Biol Res Nurs 2022; 24(2):202–215.

20. Simons RL, Lei MK, Klopack E, Beach SRH, Gibbons FX, Philibert RA. The effects of social adversity, discrimination, and health risk behaviors on the accelerated aging of African Americans: Further support for the weathering hypothesis. Soc Sci Med 2021; 282:113169. doi: 10.1016/j.socscimed.2020.113169

21. Krieger N, Chen JT, Testa C, Roux AD, Tilling K, Watkins S, Simpkin AJ, Suderman M, Smith GD, De Vivo I, Waterman PD, Relton C. Use of incorrect and correct methods to account for age in studies on epigenetic accelerated aging: implications and recommendations for best practices. Am J Epidemiol 2023; 192(5):800–811.

22. Murray P. States’ Laws on Race and Color. Cincinnati, OH: Women’s Division of Christian Service, Board of Missions of the Methodist Church, 1950.

23. Gates HL. Stony the Road: Reconstruction, White Supremacy, and the Rise of Jim Crow. New York, Penguin Books, 2019

24. Burnham MA. By Hands Now Known: Jim Crow’s Legal Executioners. 1st ed. New York, NY: W.W. Norton & Co., 2022.

25. Krieger N. Measures of racism, sexism, heterosexism, and gender binarism for health equity research: from structural injustice to embodied harm-an ecosocial analysis. Annu Rev Public Health 2020; 41:37–62.

26. Krieger N, Waterman PD, Gryparis A, Coull BA. Black carbon exposure, socioeconomic and racial/ethnic spatial polarization, and the Index of Concentration at the Extremes (ICE). Health Place 2015; 34:215–228.

27. Larrabee Sonderlund A, Charifson M, Schoenthaler A, Carson T, Williams NJ. Racialized economic segregation and health outcomes: A systematic review of studies that use the Index of Concentration at the Extremes for race, income, and their interaction. PLoS One 2022; 17(1):e0262962. doi: 10.1371/journal.pone.0262962.

28. Krieger N, Waterman PD, Kosheleva A, Chen JT, Carney DR, Smith KW, Bennett GG, Williams DR, Freeman E, Russell B, Thornhill G, Mikolowsky K, Rifkin R, Samuel L. Exposing racial discrimination: implicit & explicit measures--the My Body, My Story study of 1005 US-born black & white community health center members. PLoS One 2011;6(11):e27636. doi: 10.1371/journal.pone.0027636.

29. Krieger N, Waterman PD, Kosheleva A, Chen JT, Smith KW, Carney DR, Bennett GG, Williams DR, Thornhill G, Freeman ER. Racial discrimination & cardiovascular disease risk: my body my story study of 1005 US-born black and white community health center participants (US). PLoS One 2013; 8(10):e77174. doi: 10.1371/journal.pone.0077174.

30. Olson JL, Bild DE, Kronmai RA, Burke GL. Legacy of MESA: introduction and perspective. Glob Heart 2016; 11(3):269–274.

31. Multi-Ethnic Study of Atherosclerosis (MESA). MESA overview and protocol. https://www.mesa-nhlbi.org/aboutMESAOverviewProtocol.aspx; accessed November 28, 2023.

32. Lawlor DA, Tilling K, Davey Smith G. Triangulation in aetiological epidemiology. Int J Epidemiol 2016; 45(6):1866–1886.

33. Watkins SH, Ho K, Testa C, Falk L, Soule P, Nguyen LV, FitzGibbon S, Slack C, Chen JT, Davey Smith G, De Vivo I, Simpkin AJ, Tilling K, Waterman PD, Krieger N, Suderman M, Relton C. The impact of low input DNA on the reliability of DNA methylation as measured by the Illumina Infinium MethylationEPIC BeadChip. Epigenetics 2022; 17(13):2366–2376.

34. Min JL, Hemani G, Davey Smith G, Relton C, Suderman M. Meffil: efficient normalization and analysis of very large DNA methylation datasets. Bioinformatics 2018; 34(23):3983–3989.

35. Houseman EA, Accomando WP, Koestler DC, Christensen BC, Marsit CJ, Nelson HH, Wiencke JK, Kelsey KT. DNA methylation arrays as surrogate measures of cell mixture distribution. BMC Bioinformatics 2012 May 8;13:86. doi: 10.1186/1471-2105-13-86.

36. Liu Y, Ding J, Reynolds LM, Lohman K, Register TC, De La Fuente A, Howard TD, Hawkins GA, Cui W, Morris J, Smith SG, Barr RG, Kaufman JD, Burke GL, Post W, Shea S, McCall CE, Siscovick D, Jacobs DR Jr, Tracy RP, Herrington DM, Hoeschele I. Methylomics of gene expression in human monocytes. Hum Mol Genet 2013; 22(24):5065–5074.

37. Krieger N, Chen JT, Coull BA, Beckfield J, Kiang MV, Waterman PD. Jim Crow and premature mortality among the US Black and White population, 1960-2009: an age-period-cohort analysis. Epidemiology 2014; 25(4):494–504.

38. Caughey E, Warshaw C. The dynamics of state policy liberalism, 1936-2014. Am J Polit Sci 2016; 60(4):899–913.

39. Krieger N, Smith K, Naishadham D, Hartman C, Barbeau EM. Experiences of discrimination: validity and reliability of a self-report measure for population health research on racism and health. Soc Sci Med 2005; 61(7):1576–1596.

40. US Census Bureau. American Community Survey (ACS). https://www.census.gov/programs-surveys/acs/; accessed November 28, 2023.

41. US Census Bureau. Poverty thresholds. https://www.census.gov/data/tables/time-series/demo/income-poverty/historical-poverty-thresholds.html; accessed November 28, 2023.

42. Coull BA, Hobert JP, Ryan LM, Holmes LB. Crossed Random Effect Models for Multiple Outcomes in a Study of Teratogenesis. Journal of the American Statistical Association. 2001 Dec 1;96(456):1194–204.

43. Fiorito G, Pedron S, Ochoa-Rosales C, McCrory C, Polidoro S, Zhang Y, Dugué PA, Ratliff S, Zhao WN, McKay GJ, Costa G, Solinas MG, Harris KM, Tumino R, Grioni S, Ricceri F, Panico S, Brenner H, Schwettmann L, Waldenberger M, Matias-Garcia PR, Peters A, Hodge A, Giles GG, Schmitz LL, Levine M, Smith JA, Liu Y, Kee F, Young IS, McGuinness B, McKnight AJ, van Meurs J, Voortman T, Kenny RA; Lifepath consortium; Vineis P, Carmeli C. The role of epigenetic clocks in explaining educational inequalities in mortality: a multicohort study and meta-analysis. J Gerontol A Biol Sci Med Sci 2022; 77(9):1750–1759.

44. Dugué PA, English DR, MacInnis RJ, Jung CH, Bassett JK, FitzGerald LM, Wong EM, Joo JE, Hopper JL, Southey MC, Giles GG, Milne RL. Reliability of DNA methylation measures from dried blood spots and mononuclear cells using the HumanMethylation450k BeadArray. Sci Rep 2016; 6:30317. doi: 10.1038/srep30317.

45. Schmitz LL, Duque V. In utero exposure to the Great Depression is reflected in late-life epigenetic aging signatures. Proc Natl Acad Sci U S A 2022; 119(46):e2208530119. doi: 10.1073/pnas.2208530119.

46. Petrovic D, Carmeli C, Sandoval JL, Bodinier B, Chadeau-Hyam M, Schrempft S, Ehret G, Dhayat NA, Ponte B, Pruijm M, Vineis P, Gonseth-Nusslé S, Guessous I, McCrory C, Bochud M, Stringhini S. Life-course socioeconomic factors are associated with markers of epigenetic aging in a population-based study. Psychoneuroendocrinology 2023; 147:105976. doi: 10.1016/j.psyneuen.2022.105976.

47. Schmitz LL, Zhao W, Ratliff SM, Goodwin J, Miao J, Lu Q, Guo X, Taylor KD, Ding J, Liu Y, Levine M, Smith JA. The socioeconomic gradient in epigenetic ageing clocks: evidence from the Multi-Ethnic Study of Atherosclerosis and the Health and Retirement Study. Epigenetics 2021 Jul 6:1–23. doi: 10.1080/15592294.2021.1939479

48. Woodard K, Theoharis J, Purnell B. The Strange Careers of the Jim Crow North Segregation and Struggle Outside of the South. New York, NY: New York University Press, 2019.

49. Campisi M, Mastrangelo G, Mielżyńska-Švach D, Hoxha M, Bollati V, Baccarelli AA, Carta A, Porru S, Pavanello S. The effect of high polycyclic aromatic hydrocarbon exposure on biological aging indicators. Environ Health 2023; 22(1):27. doi: 10.1186/s12940-023-00975-y.

50. Gao X, Huang J, Cardenas A, Zhao Y, Sun Y, Wang J, Xue L, Baccarelli AA, Guo X, Zhang L, Wu S. Short-term exposure of PM_2.5_ and epigenetic aging: a quasi-experimental study. Environ Sci Technol 2022; 56(20):14690–14700.

